# Keeping care alive in 2025-26 humanitarian Somalia: routine digital data and frontline and managerial perspectives on integrated services

**DOI:** 10.64898/2026.10.30.26366902

**Authors:** Lucie Clech, Mohamed Ali Maalim, Robert Otieno Nyangah, Donald Otieno Ochieng, Mohamed Sheikh Omar Mohamud, Jihaan Hassan, Valéry Ridde

## Abstract

In conflict-affected settings facing polycrisis and a contraction in humanitarian funding, service integration helps sustain care with fewer resources, yet lacks a simple measure. We examine how the Caafimaad+ consortium’s vaccination, nutrition, and mental health services functioned under pressure in Somalia in 2025-2026. Using the Levesque conceptual framework with a sequential mixed-methods approach, we organised participatory workshops with 21 frontline workers, then nine managers, from nine structures around Mogadishu area in November-December 2025. We built an integration indicator from routine data for six structures (April 2025-March 2026). Workers described continuous pressure worsened by US funding cuts and ranked supply availability as most critical dimension. Facilities kept running through constant adaptations and informal mechanisms, especially smaller facilities. Vaccination-to-nutrition integration rate declined significantly, reflecting a dilution effect. The nutrition-to-vaccination rate showed a similar trend but non-significant. The fall in nutrition contacts matched supply shortage reports. This low-cost, reproducible approach refine service-volume monitoring where data collection is risky. Finally, our study identified a second, overlooked group: the frontline workers themselves. They sustain services through their informal efforts, but at the expense of their own well-being.

## Introduction

The humanitarian sector is experiencing a sustained funding contraction, with a 20% drop in 2025 (ALNAP 2026), forcing agencies to rethink their approach and do more with less. In increasingly complex humanitarian crises and limited resources, health and humanitarian systems must be resilient to continue serving populations (Truppa et al. 2026). The concept of resilience, defined as a system’s capacity to prepare for, absorb, and adapt to shocks while maintaining functions, has been used in health systems studies as a framework for understanding how services survive compounding pressures in fragile contexts (Truppa et al. 2024; Ridde et al. 2025). Care integration has been cited as a strategy to improve health system performance (Kozieł et al. 2023; Suter et al. 2017). For example, it coordinates patients’ care across different services, reducing missed care opportunities and easing pressure on individuals, households, overstretched resources, and health systems. For these reasons, governments, donors and coordination bodies increasingly require organisations to demonstrate service integration. In addition, because service integration requires coordination between services, it also offers a view of system resilience. Declining integration under strain can reveal coordination disruption that would remain invisible in routine service volume alone.

Yet humanitarian and health organisations lack simple, low-cost ways to know whether integration is actually happening on the ground. Global monitoring frameworks illustrate this gap. The UHC Service Coverage Index (SCI), the global indicator used to monitor universal health coverage, combines 14 indicators representing access to essential services (World Health Organization & World Bank 2023). This index does not determine whether the same individuals receive multiple services. Programmatic guidance for humanitarian services shows another limitation. The recent H3 package for humanitarian health services, developed in part from service packages used in Somalia and other fragile settings, promotes integrated approaches to service delivery, such as nutrition and communicable disease services for child health programmes, but does not specify how to measure such integration in practice (World Health Organization 2024). Adding to this difficulty, the scientific literature has no single dominant approach to measuring integration, and definitions and methods vary widely across settings. Despite repeated calls to develop more robust integration indicators (Suter et al. 2017), most existing evidence relies on qualitative or organisational assessments (staff perceptions, co-location audits, facility-level analyses) or on outcome-based impact evaluations (comparing a population with integration to one without, based on patient recall), without directly observing whether the same patients used services together, or are small-scale cross-sectional approach (Kozieł et al. 2023; Neill et al. 2023; Strandberg-Larsen et Krasnik 2009). These approaches are valuable but can be costly, slow, and rarely repeatable.

In conflict-prone areas, or whenever field access is constrained, workers’ expertise is indeed a valuable source of knowledge about how services actually function. Another type of data, routine data, may be overlooked and could complement this approach. Patient-level service data collected through routine digital systems is largely underused for integration monitoring, despite offering a simple, low-cost, and adaptable way to measure integration directly from what is already being recorded, without an additional data-collection burden. This is particularly valuable in contexts where collecting primary data is costly and can pose real risks to staff and communities, especially in hard-to-reach or conflict-affected areas. This paper addresses this methodological gap by combining participatory qualitative data with the development of a quantitative indicator of service integration based on routine digital data, in Somalia.

Somalia experienced one of the world’s most critical humanitarian crises in 2025-2026. This was driven by a polycrisis (Lawrence et al. 2024) of environmental, security, and economic shocks (UNICEF 2026b), further exacerbated by a policy-driven funding crisis. This crisis largely stemmed from US funding cuts which forced the closure of hundreds of health and nutrition facilities (OCHA et al. 2026; OCHA 2025), as population needs increased. While health system resilience to environmental and security shocks has sparked some recent interest, how these systems, and the integration of care within them, respond when a policy-driven shock is added to an ongoing polycrisis of compounding global and local pressures remains poorly understood. This is especially urgent for children’s nutrition and immunisation (UNICEF 2026a) as well as for health and mental health, prioritised by national policies since April 2026 and since 2019 (Federal Ministry of Health and Human Services 2019), respectively.

Using a mixed-methods approach, this paper examines how services functioned under pressure and how staff adapted to maintain care, drawing on frontline workers’ and facility managers’ perspectives in Mogadishu-area facilities and routine patient digital-level data.

## Material and Methods

This study uses a mixed-methods sequential design that combines a main qualitative participatory component followed by a complementary embedded quantitative component developed and used to corroborate, nuance or challenge the qualitative findings.

### Conceptual Framework

Data collection and analyses were structured around the CLIMHB framework on health system resilience (Ridde et al. 2025; Clech et al. 2025, 2022), combining DFID’s framework (DFID 2011) and Levesque five dimensions of health care access (approachability, acceptability, availability, affordability and appropriateness) (Levesque et al. 2013) to examine how service integration functioned under pressure.

We used Levesque’s supply-side dimensions in their original framework except for affordability. Because care is free for consortium patients, we reinterpreted affordability as monetary, non-monetary, and informal costs for staff and services. This allowed us to explore service sustainability during crises. For each dimension, workshop facilitators presented one guiding question (list in SM1). Participants discussed impacts and responses for each dimension. Some items have been found to relate to several dimensions. The final classification (Figures 1 and 2) includes adaptations to the original framework, as discussed during the workshops by the participants, and is presented in the results and in SM1.

**Figure 1:**
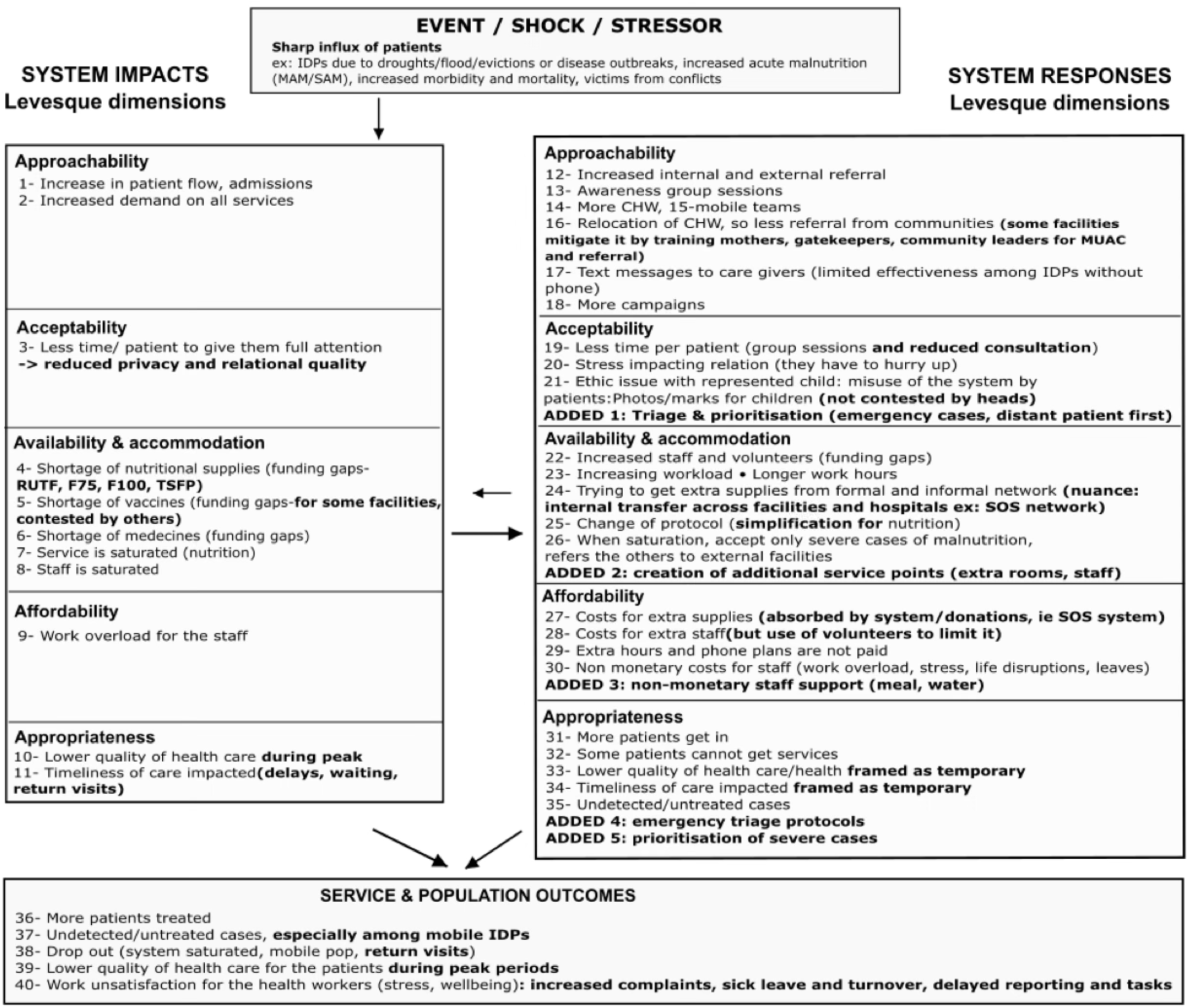
vaccination and nutrition: Impacts and responses to patient surges across Levesque’s five access dimensions, as seen and experienced by nutrition and vaccination health workers and facility heads from nine facilities in Mogadischu area. *Regular font: frontline workers’ accounts; bold: facility heads’ comments (nuances, contestations, ’ADDED’ items)*

**Figure 2:**
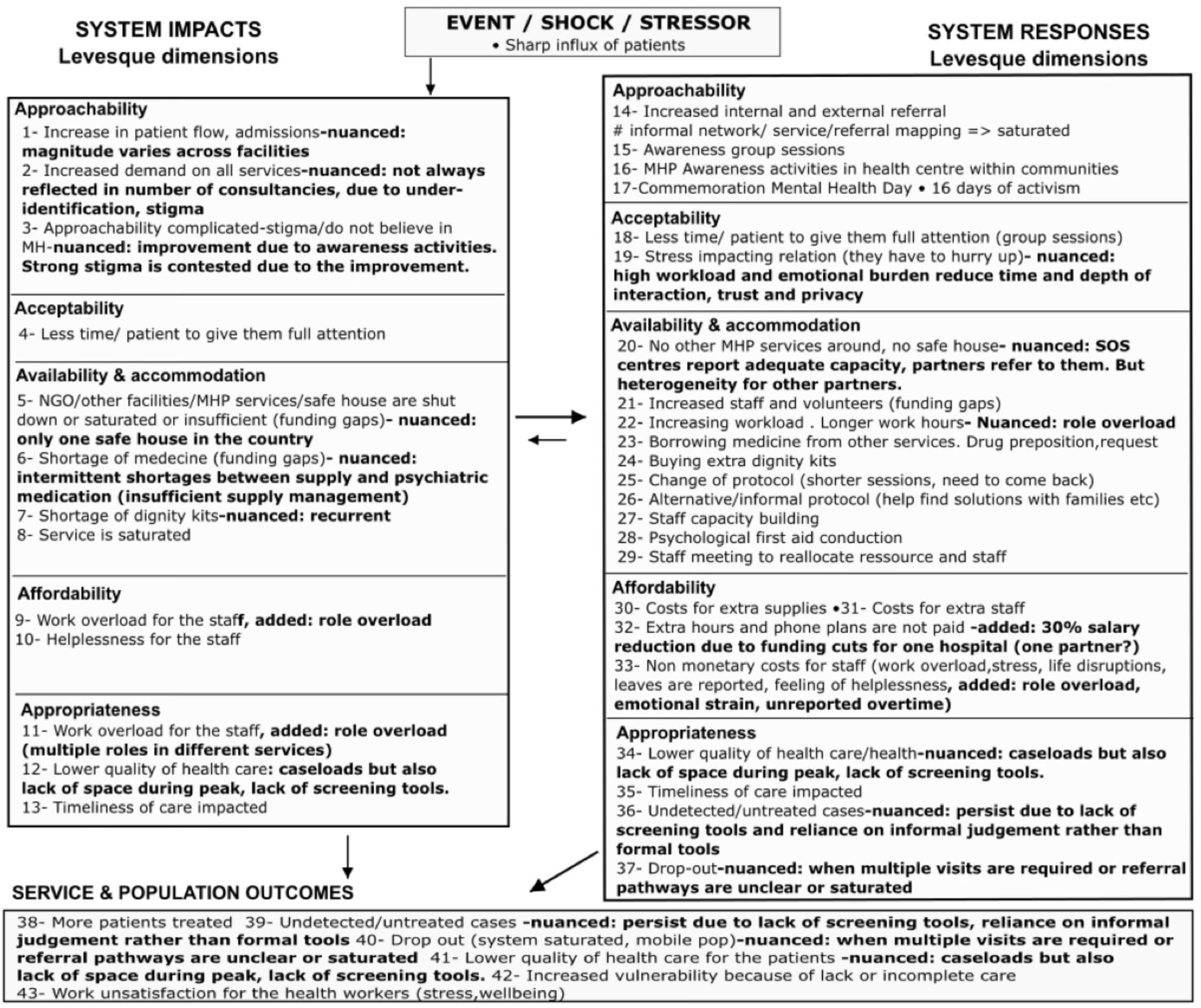
mental health and protection: Impacts and responses to patient surges across Levesque’s five access dimensions, as seen and experienced by mental health and protection workers and facility heads from nine facilities in Mogadischu area. *Regular font: frontline workers’ accounts; bold: facility heads’ comments (nuances, contestations, ’ADDED’ items)*

Setting: Somalia

Somalia faces multiple shocks, including drought, floods, insecurity, and displacement (UNICEF 2026b), further intensified by US funding cuts. These cuts led to the closure of many NGOs and health and nutrition facilities in 2025-2026 (OCHA 2025) (OCHA et al. 2026) (SM2).

These closures increased demand on the Caafimaad+ consortium (a consortium of NGOs led by ACF Somalia, including for the study period five international NGOs - ACF, SOS Children’s Villages, Concern Worldwide, International Medical Corps- and four local NGOs - Juba Foundation, Lifeline Gedo, Shabelle Community Development Organisation, and Youthlink), which provides free health, nutrition, protection and WASH services across 23 severely affected districts. Consequently, service pressures intensified.

Since 2019, national efforts have prioritised integrating mental health into primary care (Federal Ministry of Health and Human Services 2019). Since April 2026, UNICEF has prioritised childhood vaccination and nutrition services (UNICEF 2026a).

Qualitative Participatory Component

Design

A qualitative, participatory component documented how frontline workers and facility heads perceived, experienced, and responded to service pressures between January and November 2025, drawing on their emic perspectives. The design was intentionally both bottom-up and top-down within each facility. Frontline workers, closest to the communities and daily service delivery, participated in the workshops first, followed by a workshop with facility managers. Facility heads were asked to comment on, add nuance to, confirm or contest the perspectives of frontline workers. This sequential design enabled comparison of operational and managerial accounts of the same events.

### Workshop Setting and Participants

Workshops were conducted with 21 frontline health, nutrition, and protection workers and nine facility heads from the same facilities, selected with the consortium management to reflect a range of sizes and levels, and located at a reasonable distance from Mogadishu for logistical feasibility. Two workshops were organised for frontline workers: one for large/medium facilities and one for small/medium facilities. One workshop was organised with heads of facilities. All workshops focused on two themes: vaccination-nutrition and health-mental health-protection. These themes reflect the consortium’s expertise. Vaccination and nutrition are long-established areas of expertise for the consortium, while mental health-protection is a recent service under development. Thus, the workshops capture how services integrate at different stages of institutional maturity. The list and description of facilities are in SM3.

### Data collection

Frontline workers workshops used a structured sequential design of participatory activities: (1) identification of events impacting the facilities since January 2025, mapped as event-impact diagrams by team (2) prioritisation of the most significant category of events (3) rating of the event’s impact across the five dimensions of Levesque (4) mapping of formal and informal responses and coordination mechanisms between services (5) identification of barriers and facilitators. These workshop activities aimed to facilitate data classification. Figures 1 and 2 summarise results from all activities.

The facility heads’ workshop reviewed and discussed the final findings (Fig. 1 and 2), generating a comparison of frontline versus managerial perspectives. Workshops were organised in English and facilitated in both English and Somali, documented through photographed outputs (lists, mind maps, tables) and notes, debriefs were audio-recorded.

### Analyses

#### Workshops

For frontline workshops, data were analysed by group (small/large facilities), theme (vaccination-nutrition and health-mental health/protection) and activity. The information was summarised and classified within the framework as events, immediate contextual effects, health system impacts (across the five dimensions of Levesque), health system responses (across the five dimensions of Levesque), and service and population outcomes, with feedback loops identified whenever possible. For the facility heads’ workshop, contrasts and nuances with frontline workers were identified.

### Quantitative Component

#### Digitalised routine data

Caafimaad+ began organising the digitalisation of routine data in late 2024 for some facilities, strengthened training, and expanded digitalisation coverage in early 2025, and by May 2026 was running in 41 consortium facilities. The Caafimaad Plus Data Digitalisation system (C+DDS) is an electronic medical record developed by the consortium, linked to a pharmaceutical inventory system and to DHIS2 for national reporting. Of the nine facilities represented during the workshops, six use the C+DDS, one uses another pre-existing system, and two are not yet digitalised because of remaining challenges such as dual paper/digital reporting during the transition, unreliable connectivity, and power interruptions. We used data from April 2025 to March 2026, starting when staff were sufficiently trained, allowing us to explore how integration evolved over time following the recent US funding cuts. Digitalised data were available for vaccination and nutrition, but not for mental health and protection. As such, the nested quantitative component focused only on integrating nutrition and vaccination, specifically for the most vulnerable children (under 1000 days).

#### Development of the Integration Indicator

Heuristic rules and heuristic matching algorithm are presented in SM4. Contacts were matched using greedy nearest neighbour matching without replacement (Austin 2014). Digitalised records captured access to service types (multiple service types might be recorded for the same consultation) and were transformed into unique contacts per day per patient for nutrition, or vaccination (or other services not considered for this study). Thus, a contact is the patient’s access to any nutrition or vaccination services on a given day or period.

We assessed integration separately in two directions: EPI-to-Nutrition (EPI→N) and Nutrition-to-EPI (N→EPI), because the two pathways involve different clinical logic and operationalisation.

From match contacts, integration rates where then calculated by 14-days periods:

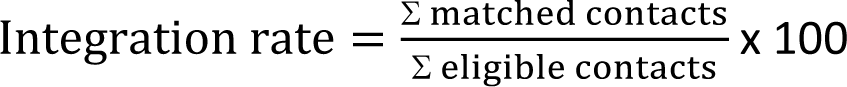

14-day periods were defined as 14-day windows starting on the Monday of each fortnight, with the first period covering 1–13 April 2025. The last incomplete period was excluded from all trend analyses. The integration indicator measures a child’s utilisation of both vaccination and nutrition services within a specific time window (same day or within 7 days). We interpret this as an indicator of integrated and continuous care, as part of Levesque’s appropriateness dimension. Utilisation is one step of Levesque framework, the point where access is realised through use of care, which supports using service utilisation as an indicator of continuity of care.

### Analyses

Descriptive analyses were conducted for the full sample of six facilities over April 2025-March 2026. Disaggregation by facilities is presented in (SM5). For trend and changepoint analysis, 14-day time series were preferred over monthly or weekly series because they provide 18 periods, each meeting at least both 5 matched contacts and 5 non-matched contacts for the overall sample. The threshold of 5 reflects a pragmatic minimum count. A higher threshold excludes more periods, and a lower threshold includes more unstable results; we also tested thresholds of 3 and 10. We applied Mann-Kendall trend tests to 14-day integration-rate series for the full sample and to integration rates in both directions. We provided Pettitt changepoint analyses for data exploration. Changepoint analysis identifies the point where the statistical characteristics of the period before and after change the most and may not reflect a discrete, datable event, but rather a gradual switch. Trend and changepoint analyses were performed on the pooled sample because matched contact counts were too small at several facilities to yield stable rate estimates. With a small numerator, a single matched contact (or non-matched) can shift the integration rate by several percentage points. Rates calculated on small samples are sensitive to sampling variability rather than reflecting real changes due to operational change or service disruption events. Statistical significance was set at p < 0.05. Rstudio software was used (Version 2023.03.1 + 446 (2023.03.1 + 446).

### Data Integration

Quantitative data were used to corroborate, nuance or challenge qualitative accounts rather than to measure the same constructs. The quantitative component is nested within the qualitative one, as it focuses on a subsample. Participants’ accounts on vaccination and nutrition integration, and on mental health integration, covered all patients at the nine facilities from January 2025, while quantitative data covered vaccination and nutrition for children under 1000 days in six of the nine facilities from March 2025.

### Ethics

Participants formally agreed to participate in these research activities. Workshop participants and quantitative patient data were anonymised. The study received ethical approval from the Ministry of Health and Human Services, Federal Republic of Somalia, through the AIM Working Group Research and Ethical Committee (Reference No: Ref/MOHHS/0257/November 2025 approved the 28^th^ of October 2025).

## Results

Qualitative participatory component

Both frontline and facility heads described a system operating routinely under cumulative, successive or continuous pressures rather than isolated crises. Figures 1 on vaccination and nutrition and Figure 2 on mental health show the classification discussed with frontline workers and facility heads.

Events cited as disrupting services from January to November 2025 included displacements driven by conflict, drought, and evictions; disease outbreaks (i.e., measles); and the introduction of new services, such as family planning and new immunisation activities. PCV (pneumonia) and rotavirus (diarrhoea) vaccines were introduced into routine immunisation in April 2025, alongside a measles catch-up campaign and subsequent polio and integrated campaigns (led by the Ministry of Health with support from WHO, UNICEF and Gavi) (SM2 for detailed chronology). We focused on displacement-related events, which most often led to a surge in patient demand and the need for immediate system scale-up (Figures 1 and 2, stressor box), because participants identified them as the primary driver of pressure.

“Funding gaps” was the terminology used by workers to describe funding issues (I.e., funding cuts (such as USAID funding cuts), delays, shortfalls, project-based funding, etc.). US funding cuts appeared as an underlying pressure rather than a single, dateable shock. Participants directly linked them to the closure of some services and facilities of other NGOs (Fig 2:5), shortages of nutrition supplies through the UNICEF supply chain (Fig. 1:4) and vaccination supplies (Fig. 1.5), shortages of medicines (Fig. 1:6, Fig. 2:6), job losses among health workers from closed facilities. These effects occurred alongside, and interacted with, other pressures affecting facilities.

External shocks such as drought, conflict, and evictions caused population displacement, camp overcrowding, deteriorating hygiene, disease outbreaks, and increased morbidity and mortality, thereby increasing demand for services (Fig. 1:1-2, Fig. 2: 1-2). Urban evictions brought new groups of patients to free services, as rising housing costs reduced their ability to afford healthcare.

These pressures affected service access in different ways across Levesque’s five dimensions. Both frontline workers and heads agreed on approachability for vaccination and nutrition, as services remained formally open and free of charge with nuance added by heads (Fig. 1: 3,16,19, ADDED 1, Fig 2. 1-3, 19). Divergence focused mostly on availability and accommodation (whether capacity matched demand), and appropriateness (whether adapted care remained adequate, and the question of continuity). Participants identified supply availability as the most critical dimension: staff shortages could be offset by extra hours or volunteers, but stock shortages could not.

Facility responses: absorbing pressure and maintaining service continuity

In response to increased demand, staff and facilities developed strategies common to both services to maintain service integration and continuity. These included increasing work hours, often without additional remuneration (Fig.1: 23, 29, Fig.2: 22, 32); reorganising the workforces across services (Fig. 1: 22; Fig. 2: 29); creating additional service points with extra rooms and staff (Fig.1: ADDED 2); prioritising emergency cases and patients from distant locations first (Fig.1: ADDED 1) and referred patients or transferred supplies across facilities and partner networks, from or outside the consortium to other facilities (Fig.1: 12, 24, 26, Fig.2: 14). The reorganisation relied partly on volunteers (Fig.1: 22, Fig.2: 21, 29) whose numbers increased as unemployment rose following US-funding cuts.

Frontline workers and managers interpreted these adaptations differently. Both described tension between patient volume and resource depletion: accepting additional patients further reduced critical supply stocks (Fig.1: 4-6, Fig.2: 6-8) and increased pressure on staff time (Fig.1: 3, 7, 8, Fig.2: 4,8). Workers described shortened consultations (Fig.1: 19, Fig2: 18, 25), simplified protocols (Fig.1: 25, Fig.2: 25), and group sessions (Fig.1: 19, Fig.2: 15, 18) (Fig.1: 32, 33, 34, 38, 39, Fig.2: 34, 35, 37), alongside work overload and fatigue, whereas facility heads viewed them as temporary, strategic responses (Fig.1: 33, 34, 39). Reports of service quality were mixed. At the start of workshops, workers rated vaccination and nutrition as unaffected, and mental health as unaffected in small facilities but very problematic in larger facilities. Staff said they maintained quality through additional effort, but at the cost of their stress and wellbeing. However, other results suggest that care quality was probably overestimated. Several workers described a concern that they were not performing their job as well as under normal conditions (Fig.1: 33, 39, 40, Fig.2: 34, 41, 43).

The health workforce bore much of the burden of maintaining continuity. Workers relied on personal and professional networks, social skills and individual resourcefulness when formal resources were insufficient (Fig.1: 24, Fig.2: 26) and reported long hours, cancelled breaks, security risks, high responsibility, turnover, concerns about care quality and, in some cases, reduced pay (Fig.1: 29, 30, Fig.2: 32, 33). Security threats, included difficulties obtaining consent for care among patients separated from their families, adding further stress for their own safety (due to blood debt) and that of their patients when they could not secure consent. We interpret this flexibility as a double-edged feature of service delivery. Smaller facilities, described as less constrained by formalised protocols, could rely more readily on frontline workers’ judgement, relationships and initiative to adapt rapidly to changing circumstances. In larger facilities, more formalised response mechanisms, including daily coordination meetings between services, staff exchange, and established protocols, provided a more structured basis for quickly adapting to increased demand. In smaller facilities, however, greater reliance on individuals placed more of the burden of adaptation on workers and facility heads. All workers agreed that this reliance on individual responsibility came at a cost to their wellbeing, stress levels, and mental health (Fig.1: 30, 40, Fig.2: 33, 43). Facility heads corroborated these accounts, citing high staff turnover, sickness and absenteeism but also increased patient complaints.

### Vaccination and nutrition services: maintaining services under increased pressure

Both services were affected by increased patient volumes, but in different ways, affecting both availability (the physical and material capacity to absorb demand) and appropriateness (the adequacy and continuity of care) (Fig.1: 4-8, 10,11). Frontline workers reported running out of ready-to-use therapeutic food (RUTF) in November 2025 and directly attributed this shortage to USAID funding cuts affecting UNICEF, the consortium’s supplier (Fig.1: 4). In some facilities, nutrition protocols were adapted: substituted products, reduced rations, or treating severe cases only and referring moderate ones. Facility heads similarly reported nutrition supply shortages, some as early as March 2025 and described subsequent restocking as insufficient. Vaccine shortages were reported by workers in both large and small facilities, but less consistently across facilities and consortium NGOs: some reported shortages, while others did not (Fig.1: 5). Nutrition services and staff were described as saturated (Fig. 1: 7, 8). Approachability was supported through community health workers, mobile teams, awareness activities (Fig.1: 13-18).

Workers reported that simplified approaches (Fig.1:25) compromised the quality and continuity of care, as well as service integration. They also reported that consultation rhythms were not aligned across services, as nutrition consultations require more time than vaccinations and are therefore harder to sustain under the same time constraints, making integration more challenging.

Regarding acceptability, reduced consultation time and stress (Fig. 1:3, 19,20) affected privacy and relational quality. Workers also raised ethical concerns about repeated presentations of the malnourished children to obtain additional nutrition supplies (Fig.1: 21), which could lead to unnecessary duplicate vaccinations and further strain limited stocks. This created tensions around fairness and rule enforcement in a service intended to remain free and equitable, with possible effects on patient-professional relationships and wellbeing. Workers noted that resentment or conflict could arise when staff attempted to prevent the unnecessary use of limited supplies.

A few cases of vaccine refusal by certain conservative groups were reported but considered unlikely to influence integration rates at the study scale.

### Mental health and protection: indirect effects of increased pressure

Frontline workers described a surge and overload across services, including in mental health and protection services (Fig.2: 1, 2, 8). However, several heads noted that demand for MHP services was stable or varied across facilities (Fig. 2: 1,2). Heads explained this difference in perspectives by the organisation of the MHP workforce: MHP staff have dual responsibilities. They are health and nutrition workers trained in MHP and could be redeployed to vaccination, nutrition or other services when demand increases (Fig. 2: 11, 22, 29). Pressure on MHP services was described less as a sharp and overwhelming increase in MHP caseloads than as reduced staff time available for MHP activities. Both groups mentioned the stigma associated with mental health and the facility’s activities to develop community awareness (Fig.2: 3, 15-17). Facility heads reported greater approachability than frontline workers, who saw stigma as a major barrier to care.

In terms of availability and accommodation, limited space made confidential access to MHP services more difficult (also affecting appropriateness, Fig.2: 12, 34), while reducing already critical stock levels for some rare mental health treatments, leading facilities to borrow medicines from other services and facilities (Fig.2:6, 23). Workers also reported direct effects of funding gaps, including the closure of already rare mental health and protection services and facilities, which concentrated demand on the remaining ones. Heads added that only one safe house exists in the country and that centres from the SOS partner reported adequate capacity and received referrals from partners, but this was not the case for other partners (Fig.2: 5,20).

Regarding appropriateness, the absence of a clear protocol to detect psychological distress was also identified as a constraint for MHP workers, as identification often relied on their own observations or those of colleagues in other services (Fig.2: 12, 34, 36). Informal referrals between staff and informal protocols to find solutions with families played an important role in identifying and directing patients (Fig.2: 14, 26), a reliance that was again double-edged. Drop-out was linked to the need for multiple visits and to unclear or saturated pathways (Fig.2: 37). Shortened consultations and group sessions (coded under acceptability and availability, Fig.2: 18,19, 25) worsen privacy, trust and the provider-patient relationship. On affordability, workers cited unpaid phone costs for referrals, unpaid extra hours, and a 30% wage reduction at one hospital to avoid layoffs (Fig.2: 32). Service costs included extra supplies and staff (Fig. 2: 30, 31). Heads reported role overload, emotional strain and unreported overtime (Fig. 2: 9, 33). Project-based MHP funding was seen as a constraint, creating uncertainty for long-term planning, less predictability for staffing and weaker integration.

### Population and system outcomes

Across services, workers and heads reported both increased treatment (Fig.1:36, Fig.2: 38), and persistent unmet needs, including undetected cases, dropout and incomplete care particularly among mobile IDPs (Fig.1: 37-38, Fig.2: 39, 40), with increased vulnerability due to lack or incomplete care (Fig.2: 42). Staff stress and patient dissatisfaction, complaints, sick leave and turnover were also reported (Fig.1:40; Fig.2: 43). Overall, facilities maintained service delivery through repeated reorganisation, but at a cost to staff wellbeing, and to the timeliness and continuity of care. This raised the question of whether these constraints were also observable in routine data through service co-utilisation.

### Quantitative Component

The integration indicator is a proxy of the continuous nature of care, taken here as one aspect of the appropriateness dimension of Levesque’s framework.

Across the nine consortium facilities, six had routine data in the same data system for the April 2025-March 2026 period; volume an integration rates were calculated at 14-day intervals (monthly results in SM6).

Contacts volume: eligible and matched contacts

#### EPI eligible contacts

EPI-eligible contacts increased significantly (Mann-Kendall tau = 0.691, p<0.001), while matched EPI contacts showed no significant trend (tau = -0.124, p=0.400) (Figure 3). Eligible contacts rose from 135 per fortnight in early April to a peak of 654 in mid-October, consistent with vaccination campaigns reported by frontline workers. Matched contacts ranged from 5 to 41 per fortnight, peaking in April (41).

**Figure 3:**
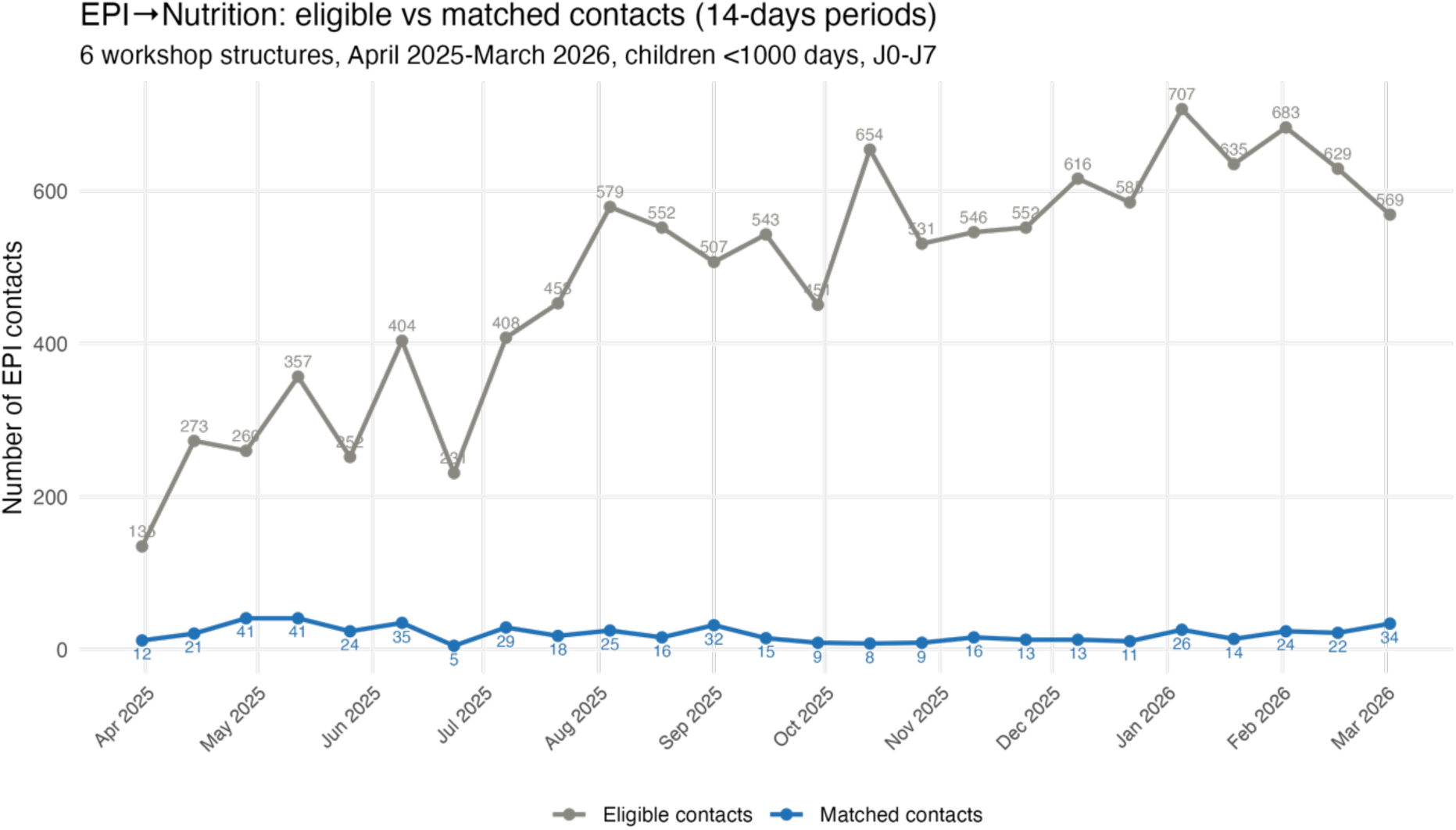
Volume of eligible and matched EPI contacts from April 2025 to March 2026

#### Nutrition eligible contacts

Eligible and matched nutrition contacts both declined significantly (eligible contacts: tau = - 0.344, p=0.017; matched contacts: tau = -0.408, p=0.005) (Figure 4). Eligible contacts peaked in the June-August period (530–768 per fortnight), then fell sharply in September-October (358–503). Matched contacts peaked in April (71 per fortnight) and declined overall, with fluctuations, including a temporary increase in September-October (up to 52 per fortnight) that did not reflect a recovery in eligible contacts.

**Figure 4:**
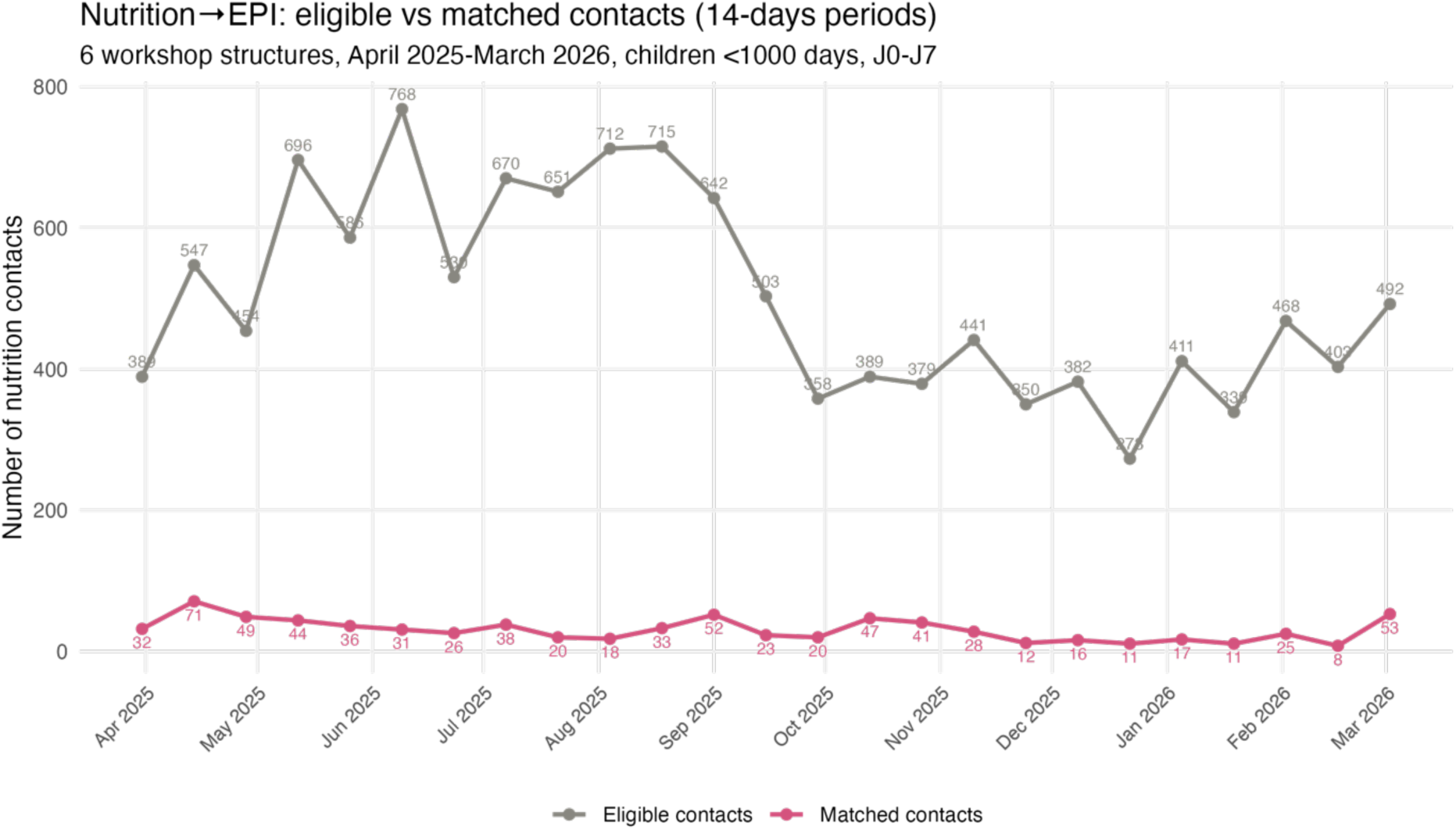
Volume of eligible and matched nutrition contacts from April 2025 to March 2026

Integration rates

Integration rates in both direction are shown in (Figure 5).

**Figure 5:**
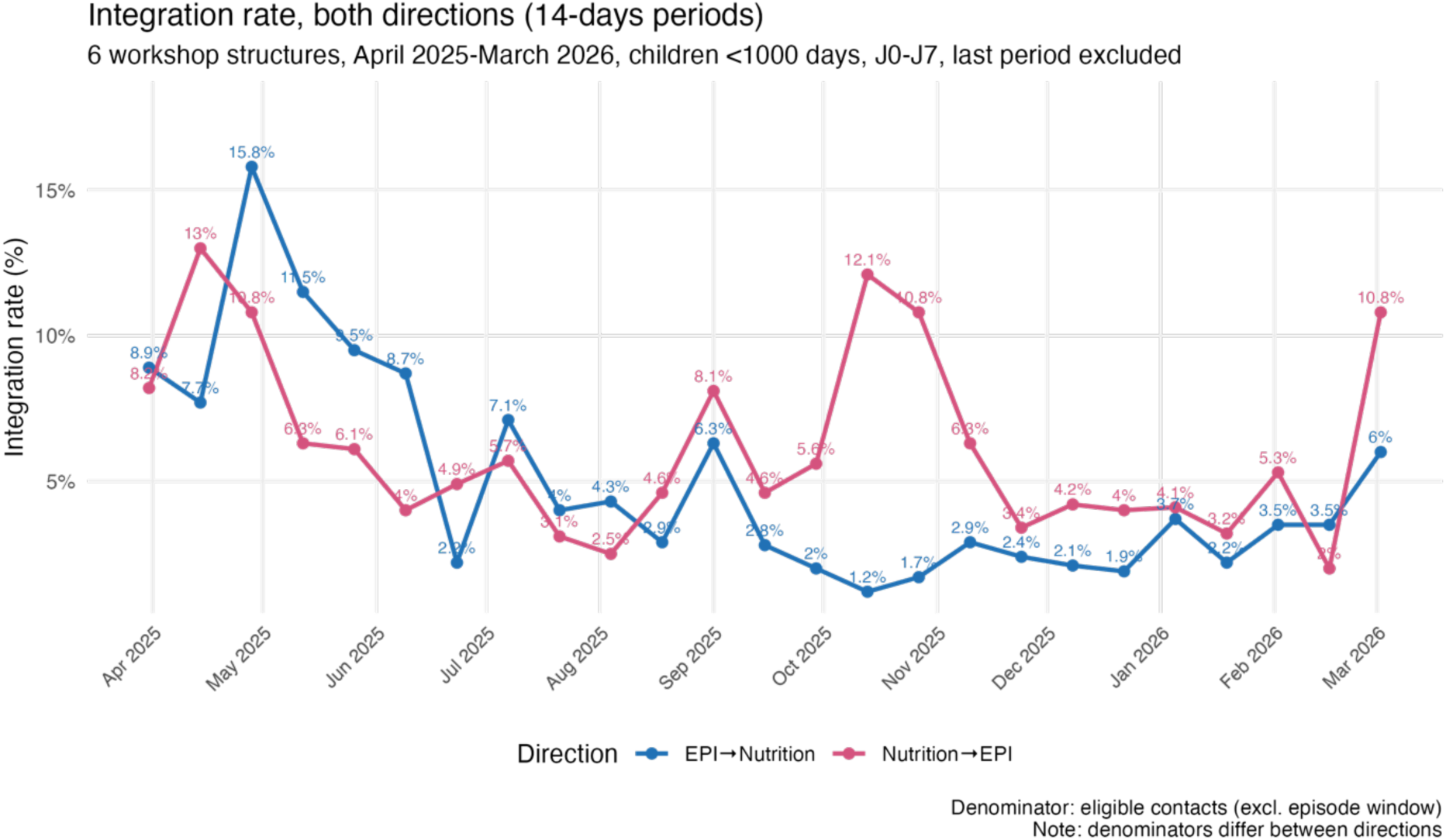
Monthly integration rates from April 2025 to March 2026

#### Trend for EPI-> nutrition integration

Integration rates from EPI to nutrition decreased significantly over the period (Mann-Kendall tau = -0.425, p = 0.003). This trend is robust with thresholds of 3 or 10 matched/non-matched contacts per point (tau = -0.425 and -0.555, respectively, p < 0.005). It partly reflects a dilution. EPI volumes increased (tau = 0.691, p < 0.001) while matched contacts showed no significant trend (tau = -0.124, p = 0.400). The growing denominator drove the rate down.

Upon arrival on site, children are ideally expected to undergo a nutrition screening first. 18.1% of matched EPI contacts (EPI-to-nutrition) were preceded by a nutrition contact within the prior seven days (22.6% within 14 days), i.e., an N-to-EPI-to-N (triple) sequence. These contacts showed a much higher co-utilisation rate (12.5%) than those without a prior nutrition contact (3.7%) within the prior seven days (12.5% vs 3.7% within 7 days; 14.3% vs 3.5% within 14 days). This probably reflects follow-up visit among children already enrolled in nutrition rather than greater cross-service referral. The decline remained significant when excluding EPI contacts preceded by a nutrition visit within 7 days (tau = -0.375, p = 0.009) and 14 days (tau = -0.341, p = 0.018)

The proportion of EPI contacts preceded by a recent nutrition contact declined from 20-25% in April 2025 to 2-4% from July 2025 onwards. This may reflect national immunisation efforts (PCV/rotavirus introduction with measles catch-up, polio rounds, Big Catch-Up for zero-dose children, SM2). As EPI volumes grew, the eligible pool increasingly included children with no recent nutrition contact or screening. Routine scale-up may have raised EPI volumes. Immunisation became the first contact after screening, and SMS reminders and defaulter tracing increased return visits. These results are consistent with frontline workers’ accounts: 1-nutrition supplies shortage (therapeutic milk in 2025 and RUTF in January 2026, and further supply delays linked to the Middle East crisis in 2026) led to modified strategies and protocols (partners kept treatment going through inter-agency loans (MSF), local preparation of F-75/F-100 (SOS), internal procurement (24000 cartons of RUTF/RUSF by SOS; Sharefood RUTF by ACF) and redistribution between sites) and fewer nutrition contacts.; 2-both services could not sustain the same rhythm during vaccination campaigns.

Overall, the decline remains consistent with the dilution effect described earlier: as EPI-eligible contacts increase, matched contacts do not follow, and the integration rate decreases.

#### Trend for nutrition-to-EPI integration

The nutrition-to-EPI rate declined, but not significantly (Mann-Kendall tau = -0.276, p = 0.058 ; threshold 10: tau = -0.220, p = 0.142). It rebounded in September-November, then decreased in December-February (Figure 5), possibly reflecting the drop in matched contacts at Lafoole HC, (peak of 45 in October, zero from December to February) and, to a lesser extent, Obosiibo Hallane HC (near-zero from August) (SM5). Balcad had almost no co-utilisation despite more EPI than nutrition contacts (n(nutrition)=363).

#### Complementary analyses: same day co-utilisations and first visits

944 same-day co-utilisation events were identified across 869 unique children (no exclusion window). Same-day pairs significantly declined (Mann-Kendall tau=-0.417, p=0.003), despite increasing EPI contacts (tau=0.600, p<0.001) and decreasing nutrition contacts (tau=-0.343, p=0.015). The proportion of EPI contacts matched with a same-day nutrition contact (matched EPI contacts with nutrition contacts in any direction/EPI contacts) declined significantly (tau=-0.550, p<0.001). The proportion of nutrition contacts matched with a same-day EPI contact (matched nutrition contacts with EPI contacts in any direction/nutrition contacts) declined but not significantly (tau=-0.181, p=0.215) (SM3), consistent with the contracting nutrition denominator partially compensating for the decline in same-day pairs.

Results remain robust when restricted to first visits of children under 1000 days. Both the number of children and the proportion of children who have access to both services in this first visit declined (Number of children: tau=-0.533, p<0.001, 14-day period time series, proportion of children: tau=-0.350, p=0.013) (SM7).

At first visit, 52.8% of children had a nutrition contact (n_epi only_= 4789, n_nutrition only_=6506, n_EPI+nutrition_=601, n _other services_ = 1570 children, April 2025-March 2026), falling from above 60% to 41% (tau = -0.464, p = 0.001).The EPI-only proportion increased significantly (tau = 0.641, p < 0.001), suggesting more new patients accessed vaccination without nutrition screening.

## Discussion

### Main findings

Three main results emerged. First, the system was described as being under continuous, cumulative pressure rather than isolated shocks. Second, frontline workers reported that the system has largely adapted through informal mechanisms, with smaller facilities relying more on individual autonomy and larger ones developing more formalised coordination mechanisms. Third, patient-level data on nutrition-vaccination integration corroborate workshop findings.

### Funding gaps, compound events and polycrisis

Somalia’s health and humanitarian sector operated in 2025-2026 in a context of polycrisis with simultaneous and interacting climate and environmental, political, and economic crises, to which US funding cuts added a cumulative pressure Workshop results suggest that US funding cuts are experienced as diffuse, cumulative pressure rather than an isolated, datable shock, which, through NGO closures and supply-chain disruptions, compounds the pressures already affecting services from population movement, disease outbreaks, organisational changes, and lack of continuous funding. Since the workshops, the Somali context has continued to deteriorate. Erratic rains during the 2025-26 Deyr season led to drought, amid severe humanitarian funding cuts, increasing the need for humanitarian food assistance. In March 2026, 3.4 million people were experiencing crisis-level hunger or worse (IPC3+)(FEWS NET 2025; World Food Program 2025). Insecurity was latent in 2025-26. In June-August 2025, Al-Shabaab regained territory lost in 2022-23, reclaiming several towns in central Somalia (Security Council Report 2025). In addition, elections triggered more instability. President Mohamud’s mandate officially ended on 15 May 2026, and a constitutional amendment extended it. Opposition contestation led to clashes between government forces and opposition fighters in Mogadishu in June 2026 (Security Council Report 2026). Conflicts in the Middle East have continued to affect international gas prices and drive up local prices of goods (World Food Program 2026a). More recently, specialists raised concerns that a severe El Niño event could affect Somalia’s environment in late 2026 (World Food Program 2026b) Finally, in 2026, the US’s decision to end financial and logistical support for the United Nations Support Office in Somalia may change the local security landscape and increase population displacement (Ismael 2026).

Taken together, these successive and overlapping developments show how compound events and polycrisis create sustained, non-linear pressure that defies the simple logic of discrete shock-recovery cycles, with each layer adding to the last.

### Triangulation of workshop and routine data

Patient-level data suggest the decline in service integration is consistent with a gradual deterioration. Access to supplies declined gradually. Facilities were partly restocked several times before stocks were depleted and the changepoint in June 2025 cannot be attributed to a unique event, it may reflect a gradual shift.

The asymmetry between the two integration directions supports this finding. EPI-to-nutrition declined through dilution, with vaccination volumes growing while matched contacts remained stable. Nutrition contacts declined : contacts with 7-day exclusion windows declined sharply between August and October 2025, and similar patterns were noticeable for same-day contacts (all contacts) and same first-day contacts.

The share of children with a nutrition service at their first visit fell from above 60% to 41% and the proportion of access to both services at that visit declined (Complementary analyses, SM7). These results cannot determine whether this reflects reduced access, a registration gap or both during patient surges. Nutrition-to-EPI contracted in both nutrition volume and matched (nutrition to EPI) contacts, reflecting supply shortages, with a resulting decreasing, near-significant tendency, and consistent results across different analyses. The asymmetry may be partly mechanical. Given the volume imbalance between the two services, nutrition contacts are more likely than EPI contacts to fall within seven days of a contact in the other service. This should not be interpreted only as a clinical gradient. Part of the decline coincide also with sharp drops in matched contact at two facilities (Lafoole, Obosiibo Hallane). In addition, frontline workers explicitly reported that nutrition teams, during patient surges, struggle to keep up with the faster pace of the EPI team, suggesting the need for extra labour force in nutrition in times of surge.

Triangulating both qualitative and quantitative data helps clarify the situation by offering a partial compromise between divergent perspectives. Some facility heads described a norm made of multiple manageable shocks, with short-term system overloads and periods of adaptation as part of a recovery from shocks. Others confirmed that facilities deal with high staff turnover, staff and patient complaints, sick leave, and staff retention issues, aligning partly with accounts from frontline workers, who reported stress and costs related to their personal lives and mental wellbeing, suggesting a complex and stressful context of adaptations. These discrepancies may reflect the structural gap between frontline workers and managerial perception, that may widen under pressure.

### Conceptual challenge of service access

This study used the Levesque framework of service access to cover the supply side of service access from the perspectives and experiences of health and nutrition professionals, with some adaptations (Levesque et al. 2013). Consortium facilities offer free care (consultations, medicines, and ambulance) to their patients, impoverished rural and urban populations, whether or not IDPs. Because we did not interview patients, we included monetary and non-monetary, formal and informal costs for professionals in the affordability dimensions. This is a conceptual shift from costs to patients to costs to professionals and services. However, this choice also indirectly connects to patients’ access to services, as costs borne by services and professionals may affect service delivery to patients and other dimensions of the framework (appropriateness, acceptability, availability etc.).

This connection was particularly clear for MHP, where affordability (our definition), availability, and appropriateness were closely intertwined. Facility heads described project-based funding that constrained planning and integration, linking service costs to gaps in care pathways. More broadly, frontline and head accounts differed, or were nuanced, across four of the five dimensions for MHP: approachability, availability, affordability, and appropriateness. This cross-dimensional divergence also reflects the recent introduction of MHP services and their ongoing organisation.

Patient-level data offer another example for vaccination and nutrition. Routine data show how pressure on EPI services reduces appropriateness for patients: the EPI-to-nutrition rate declined, partly reflecting national vaccination campaigns, changes in patient-flow organisation, and service pressure. The pattern reflects how services were organised and resourced as much as staff being overwhelmed, and both readings fit frontline workers’ accounts.

Finally, Levesque dimensions were easier to apply from a service-delivery perspective than from patients’ access experience, which suits worker expertise but may have led them to overestimate the quality of care. This is unsurprising, as most questions in tools built on Levesque’s framework focus on patients (Cu et al. 2021) rather than providers, so using it with health workers may require reinterpreting dimensions from their standpoint.

## Health System resilience

Workers from smaller facilities tended to report more informal responses to shocks than workers from larger ones. According to Blau (1970), a structure’s size predicts its formalisation because of a trade-off: the more staff there are, the more costly it is to maintain informal coordination, as each additional member increases the volume of information to process. Protocols reduce this burden by establishing roles and decision-making rules in advance, but at the expense of the flexibility informal coordination offers in unstable contexts (Burns et Stalker 1994).

This also aligns with network theory, as staff develop formal and informal connections inside and outside facilities. Granovetter’s foundational work (1973) suggests that novel information often travels through weak ties rather than strong ones. Adapted to our context, the informal ties here, connections outside formal structures, or personal connections outside of formal protocols (i.e. in other services) may play a similar role in carrying information and helping find solutions that more formal channels may not transmit. Thus, some degree of informal or semi-formal coordination remained necessary even in larger structures to allow room for adjustment. This is consistent with empirical evidence from US hospitals during COVID-19, which shows that effective adaptation depended on hybrid arrangements: formal mechanisms provided coordination and accountability, while informal and semi-formal mechanisms preserved room for adjustment (Hayirli et al. 2024).

This contrasts with a comparable study of primary healthcare centres in central Mali in context of armed conflicts, where no formalised response was observed, regardless of facility size (Lerosier et al. 2023). One hypothesis is that different institutional histories may explain this result: while the Malian health system experienced a sudden security crisis in 2012, the Somali health and humanitarian sector has functioned in a crisis context for more than three decades. This raises a question for future research: how does prolonged crisis exposure shape institutional adaptation and resilience, and does it produce different health-system responses to subsequent shocks? The duration and chronicity of crisis exposure may constitute an important dimension of resilience, alongside the intensity of shock itself. This distinction could help us distinguish resilience developed in response to sudden crises from resilience built over time through repeated exposure to crisis.

An underlying finding of this study is that frontline workers are an unrecognised population on whom service resilience depends structurally because of their informal effort. As discussed above, autonomy enables rapid adaptation but shifts risk and burden onto individuals. In other words, staff absorb part of this cost through long hours, personal risk, and emotional strain, and could benefit from mental health and protection support themselves. As organisational resilience is achieved at the expense of worker resilience, as in Tigray, Ethiopia (Tequare et al. 2026), further research is needed on humanitarian workers’ resilience, examining how repeated exposure to stress, risk, and high workload affects their wellbeing and their capacity to sustain adaptive practices over time.

In short, integration under constraint leaves little margin. Here, staff covered several roles. Stock was low. Informal and unpaid effort (extra hours, volunteers) filled the gap. Focusing on the optimisation or maximisation of resource use can reduce resilience (Almeida 2024). In unstable settings, optimisation makes systems fragile, whereas robustness keeps them working (Hamant 2023).

Resilience frameworks use different categories of processes and outcomes of resilience. For example, the DFID framework uses four categories (DFID 2011): bounce back better, bounce, recover from worst and collapse, while the ClimHB use an adapted version of DFID framework with five outcomes for the system: collapse, deteriorate, recover, improve and change (Ridde et al. 2025; Clech et al. 2022). While some facility heads describe a typical recovery process for their own facility, asking about resilience outcomes over a short period versus a longer one may offer additional insights. This also raises a question not only of time scale, but also of unit of analysis: the service, the facility, and the system, represented here by aggregated quantitative data for the six facilities on service and inter-service (with integration data). Both ClimHB and DFID frameworks characterise resilience processes and outcomes, but they do not disaggregate them by scale. Lerosier proposed an adaptation of Blanchet resilience framework for their study in Mali and distinguished three dimensions: absorb, adapt and transform (Lerosier et al. 2023; Blanchet et al. 2017), where absorb is the individual scale, adapt is the collective scale, and transform is the structural scale (or the ability of the system to evolve). In our context, and considering both the facility and consortium scales, individual-level and informal responses constitute the absorb dimension, while team reorganisation, staff meetings, triage, and protocol changes, decisions made collectively at the facility or service level, correspond to the adapt dimension. Decisions made at consortium level, such as reconfiguring supply chains and securing new types of funding, which are structural responses, fall under the transform dimension. This point was not developed in the workshop.

The consortium began adapting within existing resources from July 2025, after the USAID stop-work order, then formalised and intensified these measures from March 2026, following new multi-year funding from FCDO and renewed financing from ECHO. They cover four areas, targeting operational and institutional gaps. First, community-based financing through Health Facility Committee-managed operational funds and voluntary community co-contributions of cash, labour, fuel, and community health worker stipends to reduce dependence on any single funding source and build a locally anchored financing floor. Second, anticipatory actions through a) pre-defined shock triggers linked to ICPAC and FEWS NET seasonal forecasts, b) pre-positioned health, nutrition, WASH, and protection commodities, c) community-based surveillance by trained CHWs detecting early signals of AWD, malnutrition, maternal deaths, and GBV risks before they surface in facility data, and d) standing access to outbreak response and crisis modifier funds. Third, in mental health and protection with clearer protocols, harmonised screening tools, and systematic cross-partner standardisation to address the uneven institutional practices identified across consortium facilities. Fourth, DHIS2 digitisation of facility records to improve the timeliness, completeness, and auditability of routine data, allowing systematic comparison of service outputs against expenditure and reducing the gap between frontline reality and management perception that tends to widen under sustained service pressure. These measures reflect the consortium’s ongoing engagement rather than data collected within this study.

## Limitations

First, only facility-level staff were consulted, not patients, because access is difficult in conflict settings. Views may differ from patients’ because health and nutrition workers differ from patients in social class, education, and other characteristics (Lipsky 2010; Harrell-Bond 2002). Our definition of affordability covers only service and staff costs, not patients’ journey risks or asset losses, which workers described indirectly and which the literature has already documented (Mohamud et al. 2025). Second, we quantified only appropriateness, co-utilisation does not prove referral and utilisation is only one step of the access pathway. The indicator results from parameters set a priori: any contact as the unit of analysis, a seven-day window, both directions (low rates, no reference threshold), matching to the earliest contact (not always clinically relevant), and an exclusion window (possible minor underestimation). Linkage errors in patient identifiers could split or merge pathways (Powell et al. 2025).

However, we conducted complementary analyses to test the robustness of the findings (same-day co-utilisation, first contact only, both directions together, no exclusion windows, and the nutrition-EPI-nutrition pathway). Third, nine facilities (qualitative component) and the nested six pooled facilities (quantitative component) limit representativeness and facility-level analysis. Short series and unweighted non-parametric tests do not allow regression and causal inference, which will require longer series and organisational and contextual data.

### Lessons learned, recommendations and future directions

Quantitative co-utilisation data offer a complementary perspective to frontline workers’ experience, showing how patterns of service use change when facilities are under pressure. Changes in co-utilisation can reveal shifts in patient service usage that would not be apparent from individual service volume data alone. Co-utilisation could therefore complement routine service volume indicators by providing a broader picture of how patients move across services as conditions change.

Several lessons learned and recommendations emerged directly from this work: clearer preparedness and scaling-up measures, given that scaling-up was the most frequently cited challenge by workers; strengthening monitoring and early-warning systems by combining service-saturation indicators (patient-to-staff ratio, stock-outs, overtime) with contextual data (organisational, environmental, conflict, epidemiological); a reflexive balance between autonomy and protocol-based organisation; a reflexive approach to determining an acceptable duration of service disruption; and a protocol for mental health screening and staff support. These recommendations partly overlap with some of the adaptive and structural measures the consortium began implementing from July 2025.

This methodology may be used for retrospective analyses to support reflection on system performance and identify patterns of service disruption and adaptation. Changes in co-utilisation, alongside service volumes, could indicate changing pressure on services. This approach could be further explored as part of early warning systems, with the potential to identify emerging changes in service-use patterns and enable earlier humanitarian action.

## Conclusion

This research aims to understand how vaccination, nutrition, mental health and protection services in the Caafimaad+ consortium functioned under pressure in 2025-2026 and whether patient-level data corroborated the accounts of those who delivered the care. Frontline workers and facility heads described a system holding together through continuous, cumulative, and successive pressure, rather than isolated shocks, relying mostly on informal efforts in smaller facilities and informal and more formalised coordination in larger ones. According to participants, funding cuts had a diffuse impact through increased patient flows from compounding shocks (drought, conflict, disease outbreaks), polycrisis (i.e., climate change, Middle East crisis) and the closure of other NGOs, and reduced access to supplies (disrupted nutrition supply chains). The pressure was echoed indirectly in a significant decline in EPI-to-nutrition rate over the period, used here as a proxy of the resilience of services. Methodologically, this research contributes a low-cost, reproducible, and adaptable mixed-methods approach to measuring service integration from routine patient-level data and may offer a valuable approach for monitoring in humanitarian contexts where complex crises and compounding shocks make primary data collection challenging, dangerous, and costly.

Impoverished and displaced populations remain the most visibly underserved in this context. Yet these findings also reveal a second underserved population. Frontline staff absorb, through unpaid hours, personal risk, emotional strain, and exhaustion, the gap between what a challenged system can deliver and what patients still need. Frontline workers’ resilience is what makes the services resilient. Strengthening the system’s capacity to serve Somalia’s underserved should be strengthened alongside efforts to reduce the toll this resilience takes on those who provide it.

## Data availability statement

The data supporting the findings of this study will be made available as supplementary material upon acceptance of the manuscript for publication.

## AI use

For editing

## CRediT Roles

Lucie Clech (lead author): Conceptualisation, data curation, formal analysis, Investigation, methodology, project administration, resources, software, supervision, writing-original draft, review and editing. Mohamed Ali Maalim, Dr. Robert Nyanga, Donald Otieno Ochieng, Dr. Mohamed Sheikh Omar Mohamud: resources, writing-review and editing, funding acquisition. Dr. Jihaan Hassan: investigation, writing-review and editing. Valéry Ridde: supervision, validation, writing-review and editing

## Supporting information

conceptual framework

Somalia context

facilities

ALgorithm

monthly integration rates by facilities

monthly integration rates

same day co-utilisation

first visit same day co-utilisation

## Data Availability

All data produced in the present study are available upon reasonable request to the authors

