## Supplementary material for "Keeping care alive in 2025-26 humanitarian Somalia: routine digital data and frontline and managerial perspectives on integrated services": conceptual framework

### Supplementary materials 1: Levesque framework and adaptations

We adapted the supply side of Levesque’s framework (Levesque et al. 2013) for the workshops and coding. We illustrated the dimension with one general question to facilitate communication during the workshop. Final coding followed each dimension’s conceptual definition, which sometimes extended beyond the illustrative question.

Approachability (the ability of patients to identify that a service exists and its perceived relevance to their needs): “During the event, could people identify the service and access information about the possibility of receiving care at the facility”

Acceptability (the cultural and social fit between the service and patients’values, norms and expectations): “During the event, could the consortium maintain high professional values and norms in practices?”

Availability and accommodation (the physical existence of resources in sufficient quantity and their accommodation of patients in practices): Were staff and supplies present during the event?” was assessed for staff presence, understaffing and supply availability. The question of space and accommodation was also covered. By contrast, space adequacy for confidential consultations was classified under “Appropriateness” to reflect the distinction between physical space and its suitability for private care.

Affordability. Levesque’s definition focuses on affordability for patients (the economic capacity of patients to access care and the direct and indirect costs imposed by the service). Because health and nutrition services, nutrition and medicine supplies, hospitalisation, and ambulance services are free for Consortium patients, we reinterpreted this dimension as affordability for staff and services, departing from Levesque’s framework. This allowed us to explore service sustainability during crises. We used the question: “Are the functioning costs for the services impacted by the event?” and extended it to include monetary, non-monetary and informal costs for the staff and services.

Appropriateness (technical and interpersonal adequacy, coordination, and continuity of care): “During the event, could the consortium maintain quality and timely care?”

Finally, Levesque distinguishes acceptability (cultural and social fit between care and patient’s values and norms) from appropriateness (technical and interpersonal adequacy and continuity of care) as conceptually separate dimensions. In practice, several findings reflect both dimensions. For example, reduced consultation time could be classified as appropriateness. But it increased provider and patient stress and dissatisfaction, in ways that could affect the interaction's relational and cultural sensitivity (acceptability). Shorter consultations leave less time to adapt the communication to a patient’s education, social class or ethnic background. As such, we coded this as acceptability.
