## Supplementary material for "Keeping care alive in 2025-26 humanitarian Somalia: routine digital data and frontline and managerial perspectives on integrated services": Somalia context

Supplementary materials 2: Somalia context

Somalia, the setting for this study, is a country affected by compounding global and local shocks (UNICEF 2026b) such as severe drought, floods, insecurity and indirect impacts of the war in the Middle East, which have contributed to famine, price inflation, disease outbreaks and large-scale population movements. Policy-driven funding shortages, especially after US funding cuts, have exacerbated these environmental and security shocks and reshaped the health system. By mid-2025, 300 nutrition centres had to close (OCHA 2025). By mid-2026, more than 500 health and nutrition facilities had been shut down due to the lack of funding, disease outbreaks were not controlled, and mortality risks were rising (OCHA et al. 2026). Over the same period, the number of Somalis projected to face Emergency (IPC phase 4) has tripled in less than a year, reaching 1.9 million (OCHA et al. 2026). This context has increased population needs, while health care and support services have diminished, increasing pressure on the remaining structures.

At the service level, these pressures are especially visible for children and for mental health care. Two service areas illustrate the urgent need for integrated care: child nutrition and immunisation, and mental health. In April 2026, the national health strategy formally prioritised integrating nutrition and vaccination services for children with the launch of the Somalia Community Health Strategy 2025-2029 (UNICEF 2026a). Decades of conflict and recurring drought, flood and famines have taken a heavy toll on the mental health of Somalis. No national studies have been conducted, but small-scale studies using multiple methodologies suggest a high prevalence of mental disorders (Federal Ministry of Health and Human Services 2019). Mental health integration into primary care has thus been a national policy priority since 2019 (Federal Ministry of Health and Human Services 2019), but with modest progress, due to a fragmented and under-resourced health system (Abdi et al. 2026).

### Immunisation activities in Somalia, 2025–2026 2025-2026

Vaccination campaigns and immunisation activities, 2025–2026, in chronological order:

1) February 2025: four-day nationwide polio campaign (nOPV2), door to door, targeting 2.5 million children under five in Jubaland, Hirshabelle, South West, Galmudug and Banadir. About 2.4 million children were reached across 76 districts of central and south Somalia. Led by the Federal Ministry of Health with WHO, UNICEF and the Global Polio Eradication Initiative.

2) 23 April 2025: the Ministry of Health, with Gavi, WHO and UNICEF, introduced PCV (pneumonia) and rotavirus (diarrhoea) vaccines nationwide into routine immunisation. These were Somalia's first new antigens since 2013, launched in Mogadishu. A nationwide measles catch-up campaign was launched at the same time. Because both new vaccines are given at routine EPI visits, the introduction raised the number of EPI contacts at facilities from April–May 2025 onwards.

3) May 2025: an integrated campaign combining polio (nOPV2), PCV and measles vaccines in Somaliland, Puntland, the Northeast State and South Galkayo (outside most study areas).

4) Throughout 2025: further polio rounds and the Big Catch-Up drive, which targeted about 1.5 million zero-dose children aged 1–5 years. Health workers and CHWs traced zero-dose and under-immunised children through household visits and defaulter tracing and brought them to facilities, which kept vaccination volumes high through the second half of the year.

5) 10–31 May 2026: ECHO-funded emergency measles campaign in Hudur and Wajid (Bakool). It followed an April 2026 rise in measles cases (more than 30 confirmed) amid displacement from Baidoa. It was led by ACF under Caafimaad Plus with government supervision, for children aged 6 months to 5 years. Seven outreach teams worked door to door, combining measles vaccination with malnutrition screening, deworming and vitamin A, and two isolation centres were set up. Results: 17,138 children vaccinated (156% of the 10,952 target), 16,806 given vitamin A, 7,711 dewormed, 12,118 screened (1,295 SAM and 1,857 MAM referred), and 159 measles admissions with one death. This falls after the quantitative data period (April 2025–March 2026).

Within the consortium, routine EPI follows the national schedule (BCG at birth; Penta and OPV at 6, 10 and 14 weeks; IPV; MCV1 at 9 months; MCV2 at 15 months) at fixed and outreach sites.

UNICEF. 2026a. *Somalia Launches New Community Health Strategy to Strengthen Primary Health Care and Advance Health System Resilience*. Mogadischu.

UNICEF. 2026b. « UN Agencies Warn of Worsening Hunger and Malnutrition Crisis in Somalia as Famine Risk Emerges ». *Press Release*, mai 15. https://www.unicef.org/somalia/press-releases/un-agencies-warn-worsening-hunger.
