## Supplementary figures and images for "Keeping care alive in 2025-26 humanitarian Somalia: routine digital data and frontline and managerial perspectives on integrated services"

### facilities

#### Supplementary materials 3: facilities and their description

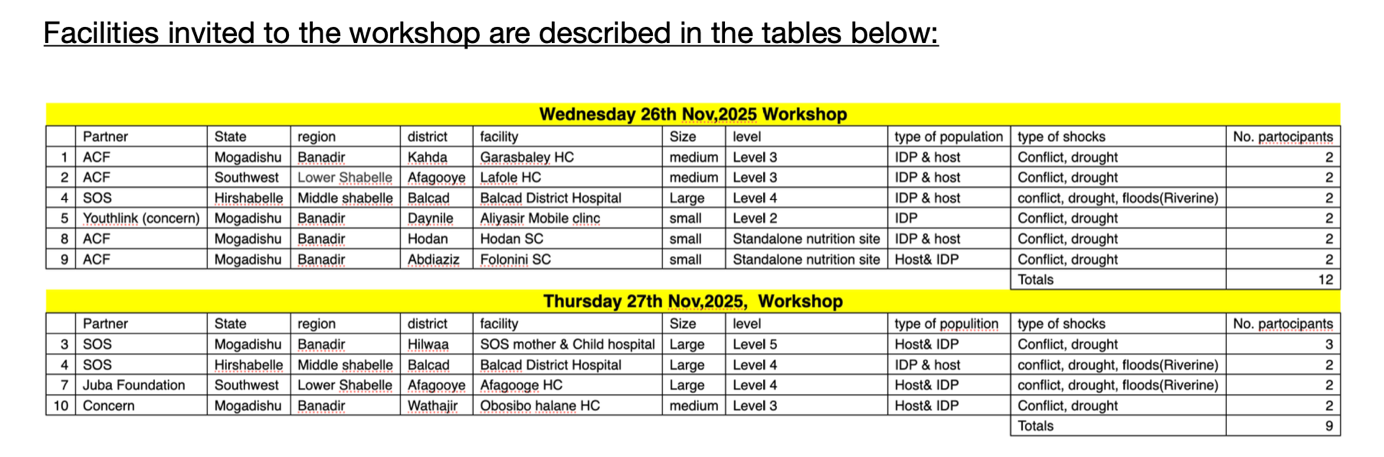
