## Supplementary material for "Keeping care alive in 2025-26 humanitarian Somalia: routine digital data and frontline and managerial perspectives on integrated services": ALgorithm

### Supplementary materials 4: heuristic matching algorithm

Each child is identified by a unique ID number, date of birth, access to health and nutrition services and date of service access. Only children under 1000 days at the time of the access were included in the analyses. We first developed heuristic rules (SM1) to define co-utilisation based on service operationalisation and local context, including integration time windows, orientation, exclusion time windows, and type of matching. Digitalised records captured access to service types (multiple service types might be recorded for the same consultation) and were transformed into unique contacts per day per patient for nutrition, or vaccination (or other services not considered for this study). Thus, a contact is the patient’s access to nutrition or vaccination services on a given day.

Integration was assessed separately in two directions: EPI-to-Nutrition (EPI→N) and Nutrition-to-EPI (N→EPI), because the two pathways involve different clinical logic and operationalisation.

The first contact is called the index; the second contact is the matching contact. For each index contact on day J, we identified the closest contact occurring between day J0 and day J+7 (both inclusive). When we found a match, we considered the pair an integrated episode starting with the index contact. Then we computed the sum of integrated (or matched) indices per month and 14-days. We calculated the health-to-nutrition integration rate as the number of matched index health contacts divided by the number of eligible health contacts. Similarly, we defined the nutrition-to-health integration rate as the number of matched index nutrition contacts divided by the number of eligible nutrition contacts. Integration rates should be interpreted as estimates of co-utilisation of services, not evidence of active referral. We then computed the number of integrated episodes per month or 14-day, when possible. Because rates can vary greatly with small sample sizes, we analysed the overall rate for the six facilities together, representing consortium context (large and small facilities together) in the Mogadishu area.

We selected the 7-day window based on three converging considerations. First, standard CMAM protocols recommend weekly follow-up visits for children with acute malnutrition, establishing the week as an operational unit for continuity of care in humanitarian nutrition programmes. Second, in cases of acute malnutrition, clinical deterioration may occur rapidly. A window longer than 7 days could therefore capture contacts that are operationally disconnected rather than reflective of genuine integrated care. Third, a 7-day period represents a pragmatic balance between clinical relevance and operational feasibility: it is sufficiently short to reflect co-utilisation, the proxy used here for possible intentional cross-referral between health and nutrition services, while also accounting for realistic scheduling constraints in fragile, conflict-affected settings such as Somalia.

The exclusion window was defined to equal the matching window (7 days), following linkage-to-care studies: once a matched pair is identified, both contacts are considered part of the same care episode, and any following index contact occurring before the matched contact plus 7 days is considered a continuation of that episode rather than a new opportunity for integration. This prevents follow-up visits happening shortly after an initial integrated contact from being counted repeatedly as separate integration opportunities. Exclusion windows apply only to matched contacts, not to eligible contacts.

For each child under 1000 days, the algorithm follows the sequence: for each index contact I (i.e. EPI contact) occurring on day t_i_:

1- If t_i_ falls within an active episode-exclusion window (opened by a previous match), the contact is not eligible and is excluded from the denominator
2- Otherwise, the contact is eligible. The algorithm searches for the earliest target contact (i.e. Nutrition contact) occurring in the interval t_m_= (t_i_, t_i_+7 days, 7 days inclusive)
3- If a target contact exists at t_m_, the index contact is classified as matched, and a new exclusion window is opened, extending from t_m_ to t_m_+ 7 days (target contact+7 days)
4- If no target contact is found, the index contact remains eligible but non-matched, and no exclusion window is opened.
5- Finally, integration rate is calculated (by month or 14-day) as:

$Integration rate=\frac{\Sigma matched contacts}{\Sigma eligible contacts}$ x 100

For each direction, integration rate is therefore defined as

$$Integration rate_{Epi to Nutrition}=\frac{\Sigma EPI matched contacts}{\Sigma EPI eligible contacts}*100$$

$$Integration rate_{Nutrition to EPI}=\frac{\Sigma Nutrition matched contacts}{\Sigma Nutrition eligible contacts}*100$$

The algorithm was piloted successfully on a small dummy sample, designed to include a series of edge cases (e.g. boundary values around the matching windows, overlapping episodes etc), to verify that it correctly classified integration episodes, before being applied to the full sample.

Heuristic rules for the contact matching algorithm

| Rule category | Rule definition |
| --- | --- |
| Unit | Contact |
| Consolidation | 1 contact per patient per day per service category |
| Window | Sliding within seven days |
| Exclusion window | Seven days following the matched contact |
| Matching | Greedy nearest per patient (closest -earliest- match to index) |
| Same-day integrated | Calculated from the timestamp |
| Direction | Unidirectional : H→N et N->H, calculated separately |
| End of period | First contacts after the 24th of last month are excluded. Rate is not calculated on the full month for the last month |
| Denominator | All health contacts (H→N) or nutrition contacts (N→H) that are not part of an ongoing integrated episode at the time of occurrence |
| Cross-facility | Integration rate with cross-facility integration included. |
