## Supplementary material for "Keeping care alive in 2025-26 humanitarian Somalia: routine digital data and frontline and managerial perspectives on integrated services": monthly integration rates by facilities

### Supplementary materials 5: Integration rates by facility/month

Presented data are monthly (and not for 14-days periods)

Eligible / matched contacts and integration rate (%), by structure and month, both directions (EN: EPI→Nutrition, NE:Nutrition→EPI), J0-J7

| Clinic | month | eligible_EN | matched_EN | rate_EN | eligible_NE | matched_NE | rate_NE |
| --- | --- | --- | --- | --- | --- | --- | --- |
| Afgoye General Hospital | 2025-04 | 44 | 1 | 2.3 | 175 | 0 | 0.0 |
| Afgoye General Hospital | 2025-05 | 24 | 0 | 0.0 | 197 | 0 | 0.0 |
| Afgoye General Hospital | 2025-06 | 20 | 1 | 5.0 | 161 | 0 | 0.0 |
| Afgoye General Hospital | 2025-07 | 80 | 1 | 1.2 | 122 | 2 | 1.6 |
| Afgoye General Hospital | 2025-08 | 358 | 9 | 2.5 | 172 | 9 | 5.2 |
| Afgoye General Hospital | 2025-09 | 274 | 5 | 1.8 | 139 | 9 | 6.5 |
| Afgoye General Hospital | 2025-10 | 358 | 0 | 0.0 | 151 | 3 | 2.0 |
| Afgoye General Hospital | 2025-11 | 462 | 8 | 1.7 | 201 | 16 | 8.0 |
| Afgoye General Hospital | 2025-12 | 543 | 6 | 1.1 | 195 | 12 | 6.2 |
| Afgoye General Hospital | 2026-01 | 554 | 12 | 2.2 | 231 | 10 | 4.3 |
| Afgoye General Hospital | 2026-02 | 525 | 15 | 2.9 | 198 | 16 | 8.1 |
| Afgoye General Hospital | 2026-03 | 393 | 4 | 1.0 | 122 | 5 | 4.1 |
| All 6 structures | 2025-04 | 450 | 40 | 8.9 | 1,003 | 108 | 10.8 |
| All 6 structures | 2025-05 | 689 | 81 | 11.8 | 1,341 | 103 | 7.7 |
| All 6 structures | 2025-06 | 681 | 56 | 8.2 | 1,373 | 68 | 5.0 |
| All 6 structures | 2025-07 | 865 | 49 | 5.7 | 1,421 | 65 | 4.6 |
| All 6 structures | 2025-08 | 1,219 | 41 | 3.4 | 1,580 | 54 | 3.4 |
| All 6 structures | 2025-09 | 1,093 | 48 | 4.4 | 1,198 | 79 | 6.6 |
| All 6 structures | 2025-10 | 1,231 | 19 | 1.5 | 801 | 72 | 9.0 |
| All 6 structures | 2025-11 | 1,207 | 31 | 2.6 | 887 | 68 | 7.7 |
| All 6 structures | 2025-12 | 1,317 | 24 | 1.8 | 765 | 29 | 3.8 |
| All 6 structures | 2026-01 | 1,436 | 44 | 3.1 | 778 | 29 | 3.7 |
| All 6 structures | 2026-02 | 1,323 | 43 | 3.3 | 883 | 33 | 3.7 |
| All 6 structures | 2026-03 | 847 | 42 | 5.0 | 755 | 67 | 8.9 |
| Alla yasir Mobile clinic | 2025-04 | 129 | 11 | 8.5 | 250 | 27 | 10.8 |
| Alla yasir Mobile clinic | 2025-05 | 187 | 8 | 4.3 | 362 | 6 | 1.7 |
| Alla yasir Mobile clinic | 2025-06 | 175 | 14 | 8.0 | 517 | 11 | 2.1 |
| Alla yasir Mobile clinic | 2025-07 | 196 | 12 | 6.1 | 456 | 8 | 1.8 |
| Alla yasir Mobile clinic | 2025-08 | 184 | 18 | 9.8 | 692 | 13 | 1.9 |
| Alla yasir Mobile clinic | 2025-09 | 164 | 11 | 6.7 | 471 | 9 | 1.9 |
| Alla yasir Mobile clinic | 2025-10 | 12 | 0 | 0.0 | 5 | 0 | 0.0 |
| Alla yasir Mobile clinic | 2025-11 | 2 | 0 | 0.0 | 6 | 0 | 0.0 |
| Alla yasir Mobile clinic | 2025-12 | 3 | 0 | 0.0 | 17 | 0 | 0.0 |
| Alla yasir Mobile clinic | 2026-01 | 62 | 3 | 4.8 | 129 | 4 | 3.1 |
| Alla yasir Mobile clinic | 2026-02 | 110 | 5 | 4.5 | 279 | 4 | 1.4 |
| Alla yasir Mobile clinic | 2026-03 | 31 | 3 | 9.7 | 300 | 4 | 1.3 |
| Balcad District Hospital | 2025-04 | 32 | 0 | 0.0 | 4 | 1 | 25.0 |
| Balcad District Hospital | 2025-05 | 15 | 0 | 0.0 |  |  |  |
| Balcad District Hospital | 2025-06 | 80 | 0 | 0.0 |  |  |  |
| Balcad District Hospital | 2025-07 | 97 | 0 | 0.0 | 44 | 0 | 0.0 |
| Balcad District Hospital | 2025-08 | 77 | 0 | 0.0 | 34 | 0 | 0.0 |
| Balcad District Hospital | 2025-09 | 118 | 0 | 0.0 | 21 | 0 | 0.0 |
| Balcad District Hospital | 2025-10 | 136 | 0 | 0.0 | 45 | 0 | 0.0 |
| Balcad District Hospital | 2025-11 | 130 | 0 | 0.0 | 76 | 1 | 1.3 |
| Balcad District Hospital | 2025-12 | 215 | 0 | 0.0 | 34 | 0 | 0.0 |
| Balcad District Hospital | 2026-01 | 226 | 0 | 0.0 | 25 | 0 | 0.0 |
| Balcad District Hospital | 2026-02 | 197 | 0 | 0.0 | 19 | 0 | 0.0 |
| Balcad District Hospital | 2026-03 | 111 | 0 | 0.0 | 51 | 1 | 2.0 |
| Garasbaley Health Center | 2025-04 | 161 | 22 | 13.7 | 188 | 48 | 25.5 |
| Garasbaley Health Center | 2025-05 | 269 | 63 | 23.4 | 222 | 21 | 9.5 |
| Garasbaley Health Center | 2025-06 | 209 | 36 | 17.2 | 197 | 10 | 5.1 |
| Garasbaley Health Center | 2025-07 | 257 | 20 | 7.8 | 318 | 6 | 1.9 |
| Garasbaley Health Center | 2025-08 | 301 | 10 | 3.3 | 257 | 7 | 2.7 |
| Garasbaley Health Center | 2025-09 | 303 | 30 | 9.9 | 279 | 41 | 14.7 |
| Garasbaley Health Center | 2025-10 | 462 | 17 | 3.7 | 352 | 17 | 4.8 |
| Garasbaley Health Center | 2025-11 | 326 | 20 | 6.1 | 341 | 5 | 1.5 |
| Garasbaley Health Center | 2025-12 | 394 | 18 | 4.6 | 361 | 17 | 4.7 |
| Garasbaley Health Center | 2026-01 | 365 | 28 | 7.7 | 354 | 15 | 4.2 |
| Garasbaley Health Center | 2026-02 | 291 | 22 | 7.6 | 288 | 13 | 4.5 |
| Garasbaley Health Center | 2026-03 | 177 | 35 | 19.8 | 246 | 57 | 23.2 |
| Lafoole HC | 2025-04 | 39 | 2 | 5.1 | 157 | 12 | 7.6 |
| Lafoole HC | 2025-05 | 109 | 1 | 0.9 | 211 | 30 | 14.2 |
| Lafoole HC | 2025-06 | 122 | 4 | 3.3 | 223 | 31 | 13.9 |
| Lafoole HC | 2025-07 | 155 | 12 | 7.7 | 244 | 34 | 13.9 |
| Lafoole HC | 2025-08 | 175 | 3 | 1.7 | 210 | 25 | 11.9 |
| Lafoole HC | 2025-09 | 110 | 1 | 0.9 | 117 | 19 | 16.2 |
| Lafoole HC | 2025-10 | 131 | 2 | 1.5 | 185 | 52 | 28.1 |
| Lafoole HC | 2025-11 | 146 | 3 | 2.1 | 195 | 45 | 23.1 |
| Lafoole HC | 2025-12 | 43 | 0 | 0.0 | 87 | 0 | 0.0 |
| Lafoole HC | 2026-01 | 74 | 1 | 1.4 | 17 | 0 | 0.0 |
| Lafoole HC | 2026-02 | 83 | 1 | 1.2 | 89 | 0 | 0.0 |
| Lafoole HC | 2026-03 | 39 | 0 | 0.0 | 29 | 0 | 0.0 |
| Obosiibo Hallane Health Center | 2025-04 | 45 | 4 | 8.9 | 229 | 20 | 8.7 |
| Obosiibo Hallane Health Center | 2025-05 | 85 | 9 | 10.6 | 349 | 46 | 13.2 |
| Obosiibo Hallane Health Center | 2025-06 | 75 | 1 | 1.3 | 275 | 16 | 5.8 |
| Obosiibo Hallane Health Center | 2025-07 | 80 | 4 | 5.0 | 237 | 15 | 6.3 |
| Obosiibo Hallane Health Center | 2025-08 | 124 | 1 | 0.8 | 215 | 0 | 0.0 |
| Obosiibo Hallane Health Center | 2025-09 | 124 | 1 | 0.8 | 171 | 1 | 0.6 |
| Obosiibo Hallane Health Center | 2025-10 | 132 | 0 | 0.0 | 63 | 0 | 0.0 |
| Obosiibo Hallane Health Center | 2025-11 | 141 | 0 | 0.0 | 68 | 1 | 1.5 |
| Obosiibo Hallane Health Center | 2025-12 | 119 | 0 | 0.0 | 71 | 0 | 0.0 |
| Obosiibo Hallane Health Center | 2026-01 | 155 | 0 | 0.0 | 22 | 0 | 0.0 |
| Obosiibo Hallane Health Center | 2026-02 | 117 | 0 | 0.0 | 10 | 0 | 0.0 |
| Obosiibo Hallane Health Center | 2026-03 | 96 | 0 | 0.0 | 7 | 0 | 0.0 |

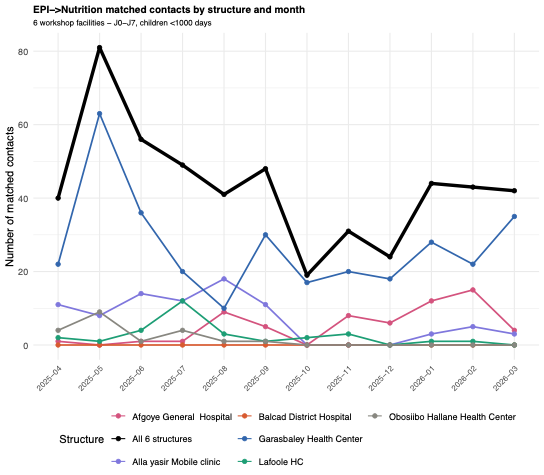

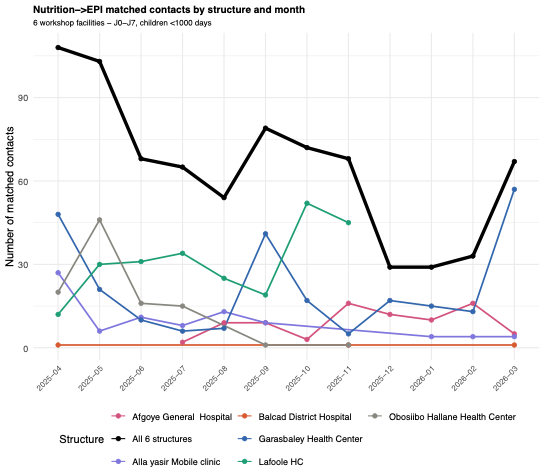
