## Supplementary material for "Keeping care alive in 2025-26 humanitarian Somalia: routine digital data and frontline and managerial perspectives on integrated services": monthly integration rates

Supplementary materials 6: Eligible, matched and integration rates per month (monthly time series, aggregated facilities)

Monthly and biweekly series are not directly comparable in terms of totals: 14-days periods do not align with calendar months, and the last incomplete biweekly period (16–24 March 2026) was excluded from trend analyses but remains included in monthly counts.

#### Nutrition eligible contacts

Over the year, both the number of eligible nutrition contacts (all nutrition contacts, except those in a 7-day exclusion window after a first contact) and the number of matched nutrition contacts (nutrition contacts followed by an EPI contact) decreased (Figure 3). Eligible contacts increased from April to August, then fell sharply in September, when matched contacts fluctuated more and did not mirror eligible contacts. For example, when eligible contacts increased (from n=1003 to n=1580) from April to August, matched contacts decreased (from n=108 to n=54). This contraction in nutrition service volume is consistent with the nutrition supply shortages reported by frontline workers and facility heads. Workshop recalls mentioned supply disruptions as early as March 2025 and insufficient restocking.

**Figure 3: Volume of eligible and matched nutrition contacts from April 2025 to March 2026**

*Value for the last month is an under-evaluation as it is calculated on 3 weeks*

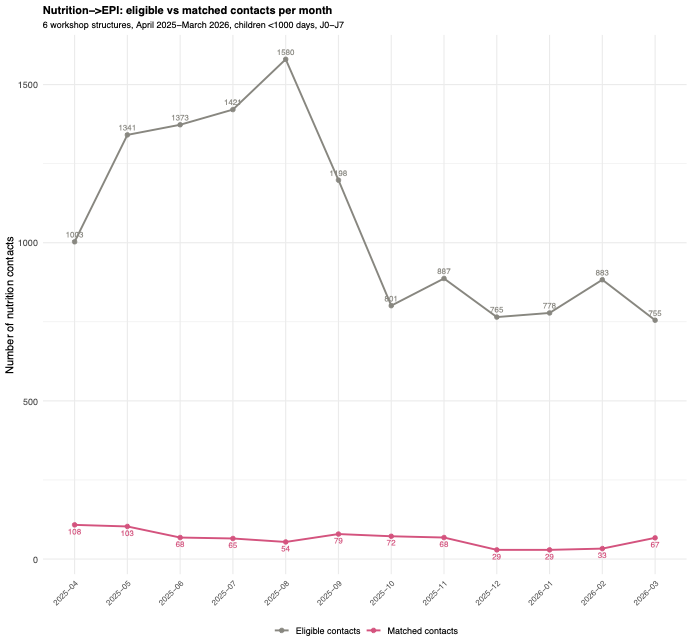

#### EPI eligible contacts

A different trend was visible for EPI contacts; it increased over the year, while the number of matched EPI contacts fluctuated. Matched EPI contacts were highest in May 2025 (n=81), when the volume of eligible EPI contacts was the third lowest for the year (n=689) (Figure 4). The matched EPI contacts were the lowest in October 2025 (n=19), when the volume of eligible EPI contacts was the fourth highest (n=1231)

**Figure 4: Volume of eligible and matched EPI contacts from April 2025 to March 2026**

*Value for the last month is an under-evaluation as it is calculated on 3 weeks*

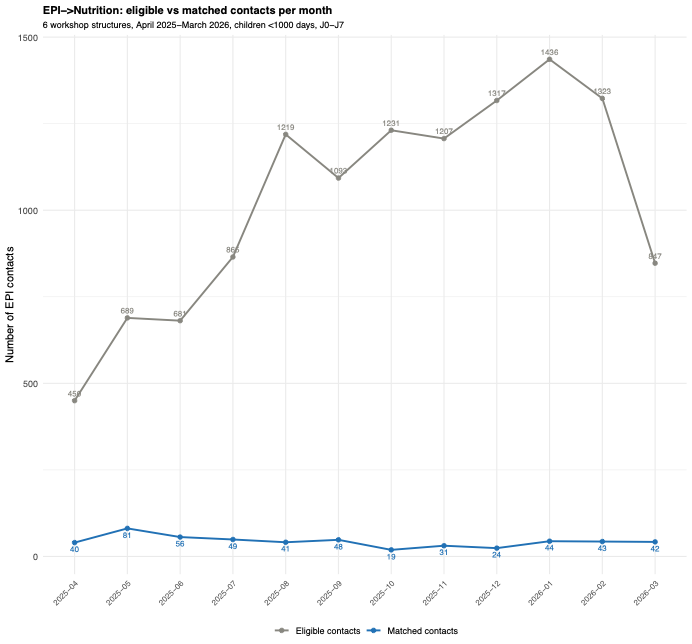

### Integration rates

In reality, patients may co-utilise services over a longer timeframe than a same-day period, and the direction of integration may provide additional information, as data show that nutrition services do not always occur during the first contact with the facility. In addition, patients may require facility services at different times of the year, not just once. When looking at eligible contacts (including first visits, other visits, and 7-day co-utilisation with an exclusion window), integration rates decreased in both directions over the year (Figure 5). Nutrition-to-EPI rebounds in September, October, and November, then decreases in December, January, and February (Figure 5). This overall decrease may relate to the sharp drop in matched contacts at Lafoole HC, (from a peak of 45 in October to zero from December to February), and to a lesser extent, at Obosiibo Hallane HC, which showed a similar pattern of near-zero matched contacts from August onward (SM2). One facilities (Balcad) recorded fewer nutrition contacts than the others (n=363 over the year). This rate is not ecplained by low nutrition contacts alone, as EPI contacts were more numerous (n=1434 eligible EPI contacts), yet almost no co-utilisation was observed.

**Figure 5: Monthly integration rates from April 2025 to March 2026**

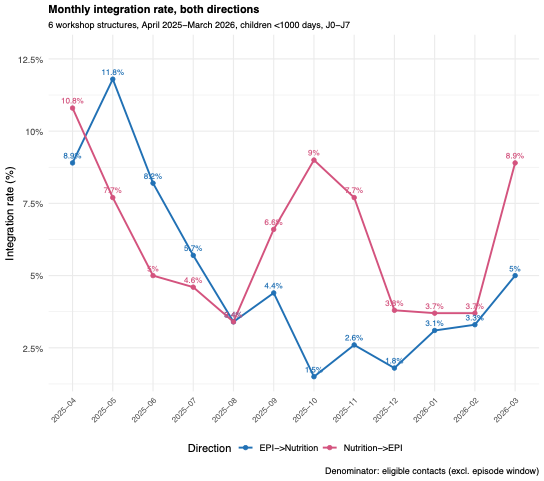

*Note: last month was calculated on three weeks only*
