## Supplementary material for "Keeping care alive in 2025-26 humanitarian Somalia: routine digital data and frontline and managerial perspectives on integrated services": same day co-utilisation

Supplementary materials 7: Co-utilisation of nutrition and vaccination (contacts) for children under 1000 days

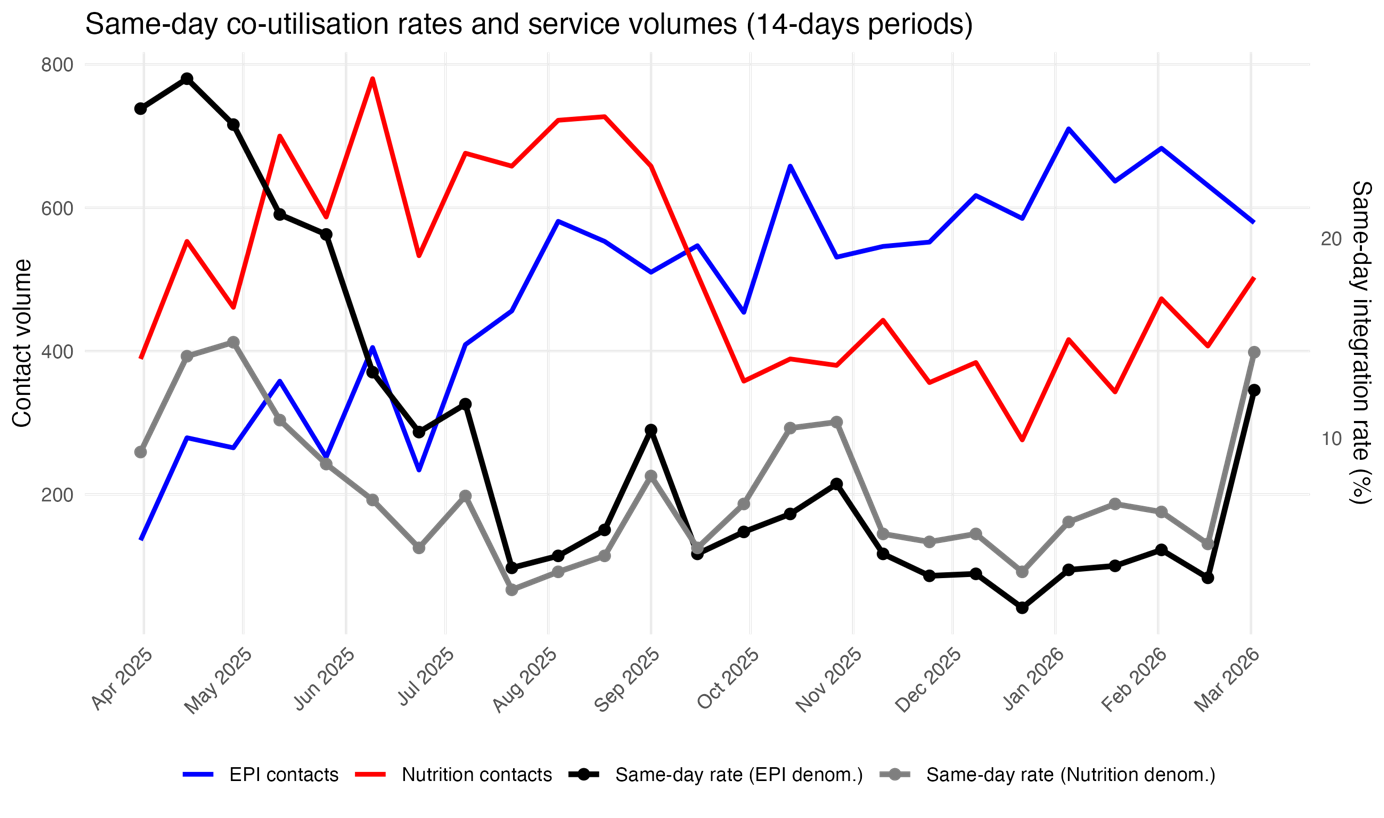
