## Supplementary material for "Keeping care alive in 2025-26 humanitarian Somalia: routine digital data and frontline and managerial perspectives on integrated services": first visit same day co-utilisation

Supplementary materials 8: co-utilisation of nutrition and vaccination for first contact for children under 1000 days

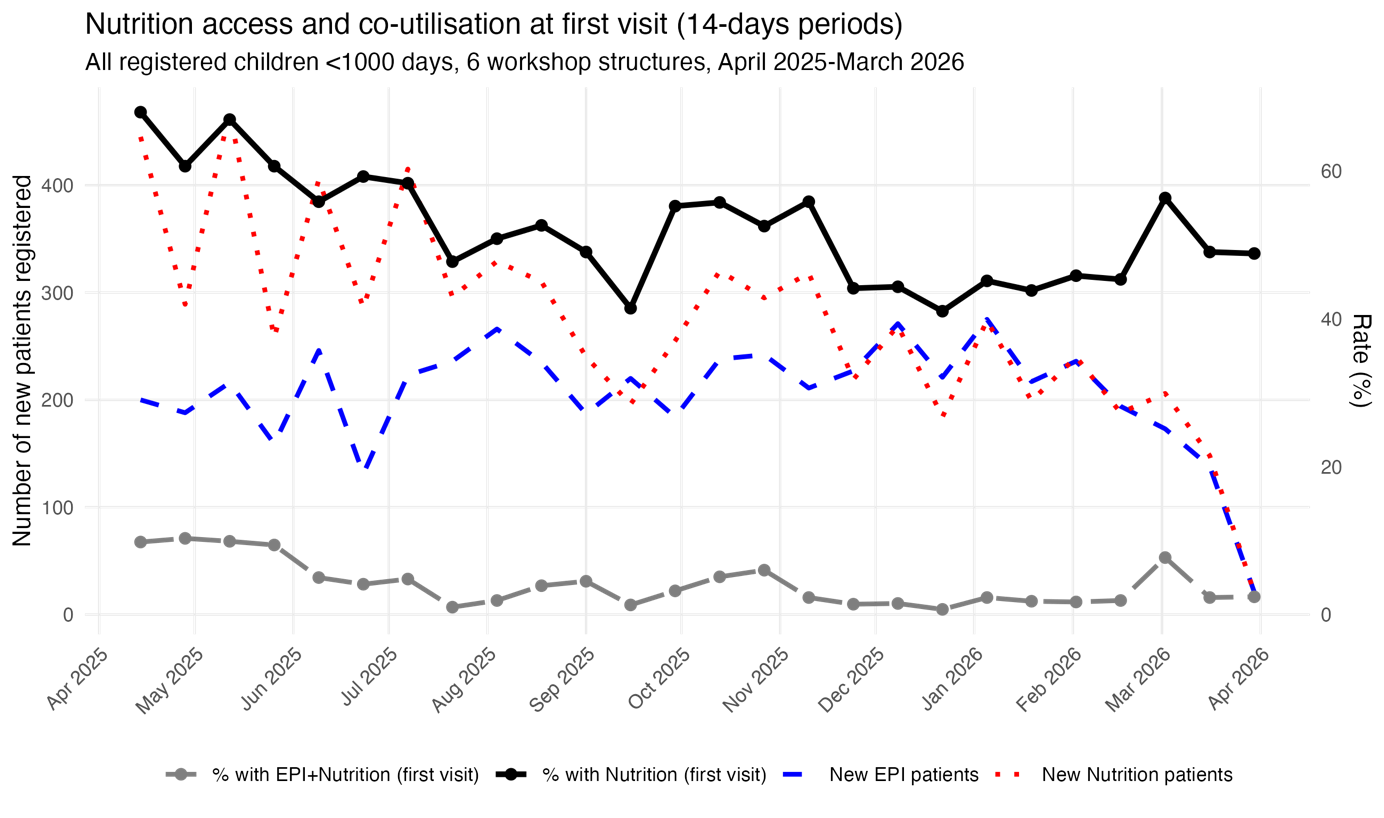
